# Impact of genome build on RNA-seq interpretation and diagnostics

**DOI:** 10.1101/2024.01.11.24301165

**Authors:** Rachel A. Ungar, Pagé C. Goddard, Tanner D. Jensen, Fabien Degalez, Kevin S. Smith, Christopher A. Jin, Undiagnosed Diseases Network, Devon E. Bonner, Jonathan A. Bernstein, Matthew T. Wheeler, Stephen B. Montgomery

## Abstract

Transcriptomics is a powerful tool for unraveling the molecular effects of genetic variants and disease diagnosis. Prior studies have demonstrated that choice of genome build impacts variant interpretation and diagnostic yield for genomic analyses. To identify the extent genome build also impacts transcriptomics analyses, we studied the effect of the hg19, hg38, and CHM13 genome builds on expression quantification and outlier detection in 386 rare disease and familial control samples from both the Undiagnosed Diseases Network (UDN) and Genomics Research to Elucidate the Genetics of Rare Disease (GREGoR) Consortium. We identified 2,800 genes with build-dependent quantification across six routinely-collected biospecimens, including 1,391 protein-coding genes and 341 known rare disease genes. We further observed multiple genes that only have detectable expression in a subset of genome builds. Finally, we characterized how genome build impacts the detection of outlier transcriptomic events. Combined, we provide a database of genes impacted by build choice, and recommend that transcriptomics-guided analyses and diagnoses are cross-referenced with these data for robustness.

## Introduction

Transcriptomics is increasingly used for studying disease etiology and diagnosis^1^. The selection of a reference genome build and corresponding genome annotation sets the foundation for the majority of transcriptome analyses, and the impact of varying annotation sources on gene expression estimates are well-documented^2–8^. However, the impact of reference genome build is less understood. Despite the release of hg19 in 2009^9^ and hg38 in 2013^10^, most academic and commercial labs align to hg19 and the majority have no plan to migrate largely due to time, computing, and staffing costs^11^. For studies that have used transcriptomics to aid rare disease diagnoses, all but two^12, 13^ published studies to date aligned to the hg19 genome build^14–27^. Additionally, release of the first ungapped human genome reference, CHM13 from the Telomere2Telomere Consortium, provides additional options for build choice and increased uncertainty regarding the impact of genome build on transcriptome analysis for diagnosis^28^. Further, despite evidence that build impacts variant calling and genetic interpretation^29–31^, and implications that reference genome build may impact RNA-seq analysis^32^, no study has systematically investigated the impact of genome build on transcriptome results.

To assess how genome build choice impacts gene quantification and the detection of outlier gene expression and splicing in a rare disease context, we conducted a comprehensive evaluation of how the hg19, hg38, and CHM13 genome builds impact RNA-seq results with gene-level resolution, and highlight rare and undiagnosed disease cases where genome build selection may affect diagnosis. Here, we significantly expanded our cohort of rare disease patients with heterogeneous disorders and their family members^15^, by generating and aligning their transcriptome data to the hg19, hg38, and CHM13 assemblies of 386 samples from 316 individuals. We identified genes with annotation-specific, differentially quantified, or build-exclusive expression across six routinely-accessed biospecimens, and assessed how these build-dependent effects influenced a transcriptome-first diagnostic interpretation. We ultimately provide a resource documenting build-dependent effects for all annotated genes, and report 2,800 genes directly impacted by build choice across a variety of scenarios (**Supplementary Figure 1)**. We expect this information to broadly enable genome build decision-making for multiple transcriptomics applications (**Supplementary Table 1)**.

## Results

### Transcriptome mapping in a rare disease cohort across genome builds

We performed RNA-sequencing for 386 samples from 316 individuals enrolled in the Undiagnosed Diseases Network (UDN) and/or GREGoR Consortium across the following biospecimens: blood, fibroblast, peripheral blood mononuclear cells (PBMCs), skeletal muscle, induced pluripotent stem cells (iPSCs), and derived neural progenitor cells (NPC) (**Figure 1a, Supplementary Table 2**). This cohort included 204 cases with a heterogenous representation of rare disease phenotypes including primarily neurological, musculoskeletal, or immune-related symptoms. Each sample was uniformly processed with a standardized pipeline and aligned to the hg19.p13, hg38.p13, and CHM13v2 genome builds (**Figure 1b, Supplementary Figure 2a**). To ensure maximally consistent gene annotations, genes were quantified using the GENCODEv35 equivalent gene annotations for each build (see Methods)(**Figure 1b**).

**Figure 1.**
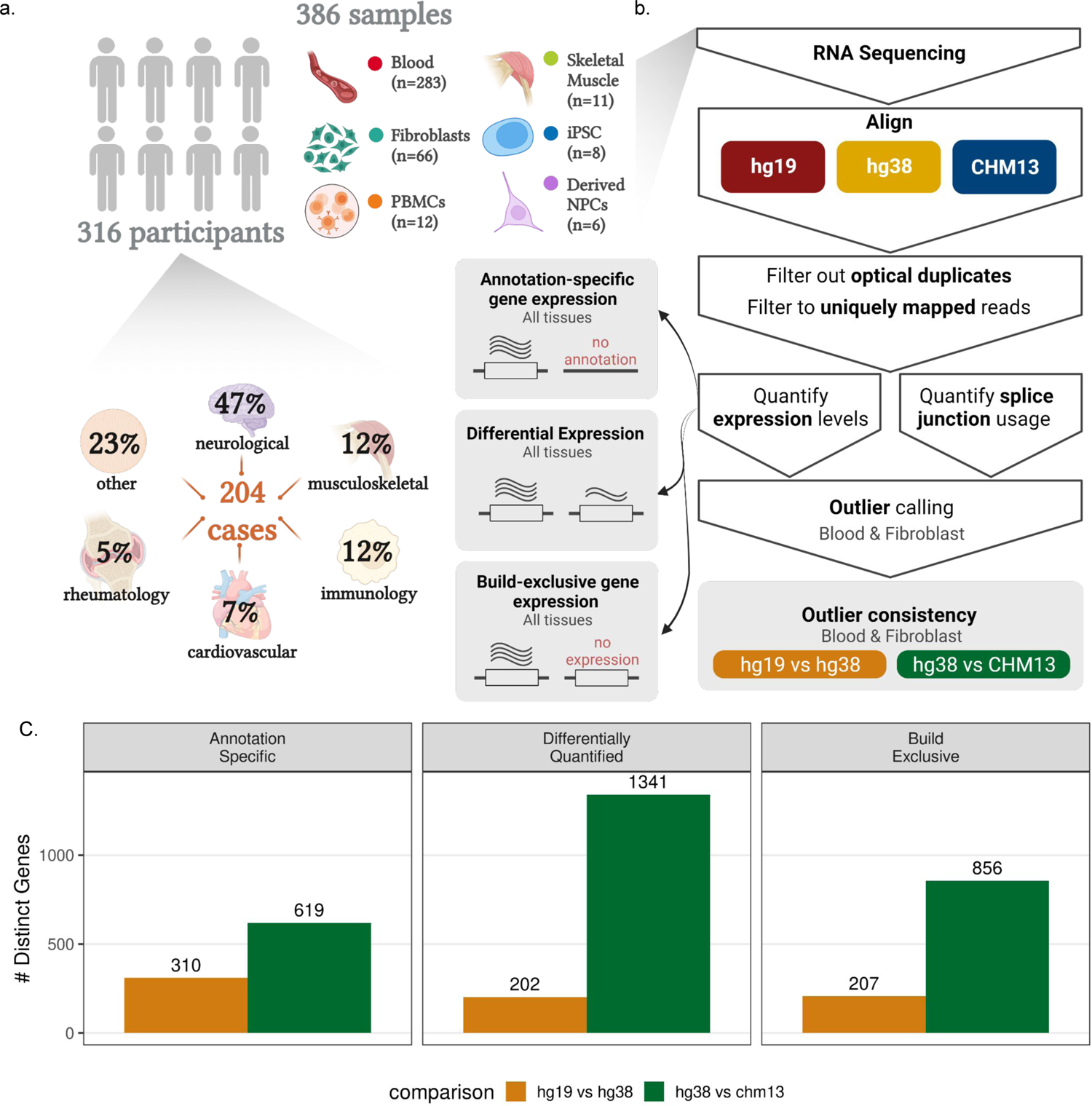
Study overview **a**, Description of cohort, including the primary diagnosis types for all probands (below) and the number of samples assayed per tissue type (right). **b,** Overview of the methodology. **c,** Barcharts displaying the total number of build-dependent events identified in the hg19 vs hg38 and hg38 vs chm13 comparisons.

Reads were uniquely aligned at a similar rate across builds (**Supplementary Figure 2b**). However, the CHM13-alignment yielded a lower proportion of multi-mapped reads and a higher proportion of unmapped reads (**Supplementary Figure 2b**). This can partially be explained by the increased proportion of unmapped reads that are classified as too short for alignment, likely due to increased complexity in the CHM13 assembly (**Supplementary Figure 2c**)^33^. The median number of non-canonical splice sites decreased by 17% between hg19 and hg38, and 18% between hg38 and CHM13, suggesting these reads are being aligned more effectively with subsequent genome builds^34^(**Supplementary Figure 2d**). Across all six biospecimens, uniquely-mapped reads quantified 78.0% of total protein-coding genes, 76.8% of Mendelian-disease-linked genes from the Online Mendelian Inheritance in Man (OMIM) database, and 64.1% of disease-associated genes from OpenTargets (score > 0.8, see **Methods**).

### Gene annotation changes across genome builds

Gene annotations describe gene structure in the context of a genomic coordinate system, and variation in how these annotations are constructed by different sources (such as GENCODE or RefSeq) can impact RNA-seq quantification^5, 7^. To minimize this annotation source bias, we leveraged the GENCODEv35-based annotation for each build with the understanding that updates to the reference genome can inherently lead to changes in the annotation. We assessed consistency of the annotations by comparing exon structure, transcript structure, and underlying genetic sequence for each gene (see **Methods, Supplementary Figure 3a**). Genes were defined as having identical gene models if the number of constituent transcripts and exons, and exon lengths were the same between builds (**Supplementary Figure 2c**). For genes with identical gene models, sequence similarity was calculated with the Jaro-Winkler similarity score which measures the minimum number of operations needed to make two sequences identical^35^. We assessed differences in annotation status, gene model, and underlying genetic sequence across 63,652 genes in hg19:hg38 and 63,710 in hg38:chm13, including 4,870 genes with a known molecular link to Mendelian diseases in OMIM annotated in at least one build across both analyses (see **Methods**).

The GENCODEv35lift37 annotation for hg19 and GENCODEv35 annotation for hg38 are largely similar, with 91.7% (58,373/63,652) of genes present in both annotations after excluding the Y chromosome. Similarly, 93.8% (59,815/63,710) of genes were present in both the GENCODEv35 annotation for hg38 and the GENCODEv35 CAT/Liftoff v2 annotation for CHM13 (**Figure 2a)**. There were 3,515 genes present in the hg19 annotation but not the hg38 annotation, and 1,764 genes present in the hg38 absent from the hg19 annotation, including five hg38-specific OMIM genes. Additionally, 322 hg38 genes (including three OMIM genes) were not mappable to CHM13 and thus not included in the CHM13 annotation, and 3,573 genes present in the CHM13 annotation had no corresponding gene model in hg38, including novel genes identified in non-syntenic regions and gene families expanded to include additional paralogs (**Figure 2a, Supplementary Figure 3a**).

**Figure 2.**
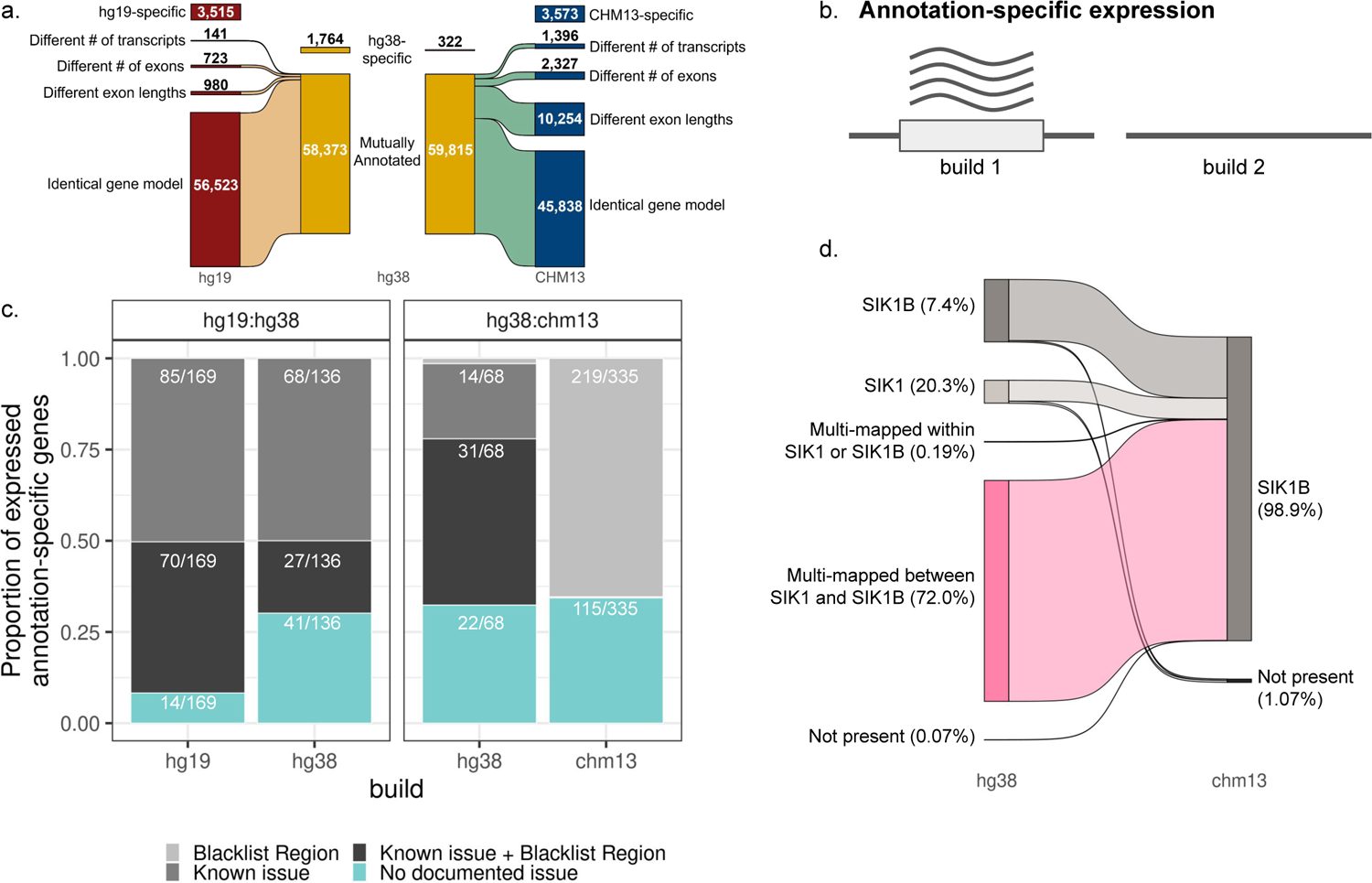
Annotation comparison identifies annotation-specific genes with detected expression **a**, Sankey diagram summarizing the number of genes that differ between the hg38 GENCODEv35 annotation and GENCODEv35lift37 for hg19 (left), and UCSC GENCODEv35 CAT/Liftoff v2 annotation for chm13 (right). **b**, Definition of annotation-specific expression events **c**, Stacked bar plots indicating how many annotation-specific genes with detected expression in at least one tissue overlap known issues in the corresponding reference genomes. Left-hand facet shows hg19-specific and hg38-specific genes from the hg19:hg38 comparison; the right-hand facet shows the hg38-specific and chm13-specific genes from the hg38:chm13 comparison. Sets containing at least 25 genes are labeled with the fraction of total expressed genes they represent within the given build and comparison. Colors represent presence (grays) or absence (light blue) of documented blacklisted regions or issues. **d**, Sankey diagram illustrating how the same RNA-seq reads are aligned to the SIK1/SIK1B locus in hg38 (left) and chm13 (right) across all samples. Percentages are based on the total number of reads aligned to SIK1 or SIK1B in either build.

Approximately 24% (13,929) of the genes present in both the hg38 and CHM13 annotations had differences in the gene model across transcript count, exon count, and exon length, compared to just 2.9% of genes annotated in both hg19 and hg38 (**Figure 2a, Supplementary Figure 3a**). Among the genes confidently linked to Mendelian disorders in OMIM and annotated in both hg38 and CHM13, 51.1% (2,490) had differences in the gene model, compared to 2.9% (142) in hg19 and hg38. An additional 30% (17,660) of genes with consistent gene models had differences in underlying genetic sequence between the hg38 and CHM13 assemblies, compared to 2.4% (1,364) of hg19:hg38 genes. Even when there were some sequence differences, they were minimal; the average Jaro-Winkler sequence similarity was quite high (mean = 0.981±0.09 hg19:hg38; mean = 0.999±0.003 hg38:CHM13 **Supplementary Figure 3b,c**). Consistency in genic exon, transcript, and sequence annotations in both build comparisons indicated that the GENCODEv35 annotation for hg38 and its liftover counterparts were sufficiently comparable, allowing us to test the effects of genome build independent of annotation for the majority of genes.

### Annotation-specific genes are quantified and can lead to erroneous results

We sought to better understand genes that were not annotated in all builds, annotation-specific genes (**Figure 2b**). We detected quantification of 169/3,515 hg19 and 136/1,794 hg38 annotation-specific genes in the hg19:hg38 comparison in at least one biospecimen, none of which were known disease genes (see **Methods**, **Supplementary Figure 4, Supplementary Table 3**). Approximately 33% of the quantified hg19 annotation-specific genes and 41% of the quantified hg38 annotation-specific genes were protein-coding or lncRNA (**Supplementary Figure 5**). Within the hg38:CHM13 comparison, we detected quantification of 68/322 hg38 and 335/3573 CHM13 annotation-specific genes, 60% and 44% of which were protein-coding or lncRNA, respectively (**Supplementary Figure 5, Supplementary Table 3**). We suggest using caution when exploring annotation-specific genes for disease associations, as the majority overlapped a blacklisted region or known assembly issue: 92% hg19, 70% hg38 (compared to hg19), 67% hg38 (compared to CHM13), 66% CHM13 (**Figure 2c, Supplementary Figure 6**).

Annotation-specific genes shed light on the importance of build selection in the context of specific disorders. For example, hg19 and hg38-expressed CFHR-Factor H complex genes CFHR1 and CFHR3 are linked with atypical hemolytic uremic syndrome^36, 37^ and fall within a region harboring population-specific copy number variations. The absence of these genes in CHM13 could be due to the reliance on a single cell line, especially in contrast to the genetic diversity from multiple cell lines underlying hg38^38^. We detected quantification of CFHR1 in fibroblast and muscle, and CFHR3 in iPS and iPS neural progenitor cells for hg19 and hg38. One study reports that detection of the disease-causing structural variants was not possible when aligning to CHM13, even with long-read sequencing^38^, suggesting CFHR-Factor H complex disorders should not be evaluated using CHM13v2. The absence of these genes in CHM13v2 likely influences mapping in CHM13 of other CFHR-Factor H complex genes – CFHR4 (also linked to atypical hemolytic uremic syndrome) is detected as quantified only in CHM13 in iPSC NPC with a median TPM of 4.4. Based on these observations, we suggest using hg38 to evaluate CFHR-Factor H complex genes.

The hg38 build contains an erroneously duplicated region, which includes paralogous genes SIK1 and SIK1B; the hg38-specific gene SIK1 has been linked to developmental and epileptic encephalopathy^39^. These two genes are identical except for a segment of the sequence encoding for a single amino acid^40^. In correcting this duplication, the CHM13 annotation used the SIK1B version of the gene. SIK1B had higher expression in CHM13 in all biospecimen types (5.5x-8.5x) compared to hg38, likely due to the removal of SIK1 which was siphoning reads in the hg38 alignment (**Figure 2d**). SIK1B is oncogenic, and further studies should be done to reveal if SIK1B is also associated with development and epileptic encephalopathy^40^. Given the transcriptomic impact of this false duplication in hg38, we suggest using CHM13 for assessment of the SIK1/SIK1B locus.

### Pairwise differential expression identifies hundreds of genes with build-dependent quantification

Genes annotated across builds may still yield differences in expression estimates between alignments; we refer to these genes as differentially quantified, as there are no true biological differences between a single sample aligned to multiple builds. To identify genes with differential quantification, we restricted to genes with at least 0.1 TPM in at least 30% of tested samples in both builds, and performed paired-sample differential expression analysis comparing gene quantifications for hg19 vs hg38 (hg19:hg38) and hg38 vs CHM13 (hg38:CHM13) using the LIMMA-DREAM framework^41, 42^.

Our selection of biospecimens allowed us to evaluate hg19:hg38 differential quantification for 31,275 genes with sufficient expression in both builds, including 72.6% (20,314/27,966) of known protein-coding and lncRNA genes (**Supplementary Table 4)**. In total, we observed 202 (128 protein-coding or lncRNA) genes with significant (Benjamini-Hochberg adjusted p-value <0.05) and substantial (abs(logFC)>1) differences in quantification between hg19 and hg38 in at least one biospecimen type (**Figure 3a,b, Supplementary Figure 6a, 7a**). Although the number of differentially quantified genes varied by biospecimen, 125 genes were consistently differentially quantified in more than one type and 29.1% (23/79) of genes tested in all six biospecimens showed significant and substantial differential quantification in all six biospecimens (**Supplementary Figure 7, 8a**). The majority of differentially quantified genes (180/202, 90%) overlapped erroneous or difficult-to-sequence regions^9, 43^ in either hg19 or hg38, with most (163/202, 81%) overlapping regions problematic in both builds (**Figure 3c**). Changes in the underlying gene sequence and/or gene model likely explain differences in expression estimates for 18 of the 22 genes that did not overlap known issues (**Figure 3c**). Taken together, known genome build errors, documented changes in the underlying build structure, and changes in gene model annotation accounted for 98% of the differentially quantified genes between hg19 and hg38 (**Figure 3c**).

**Figure 3.**
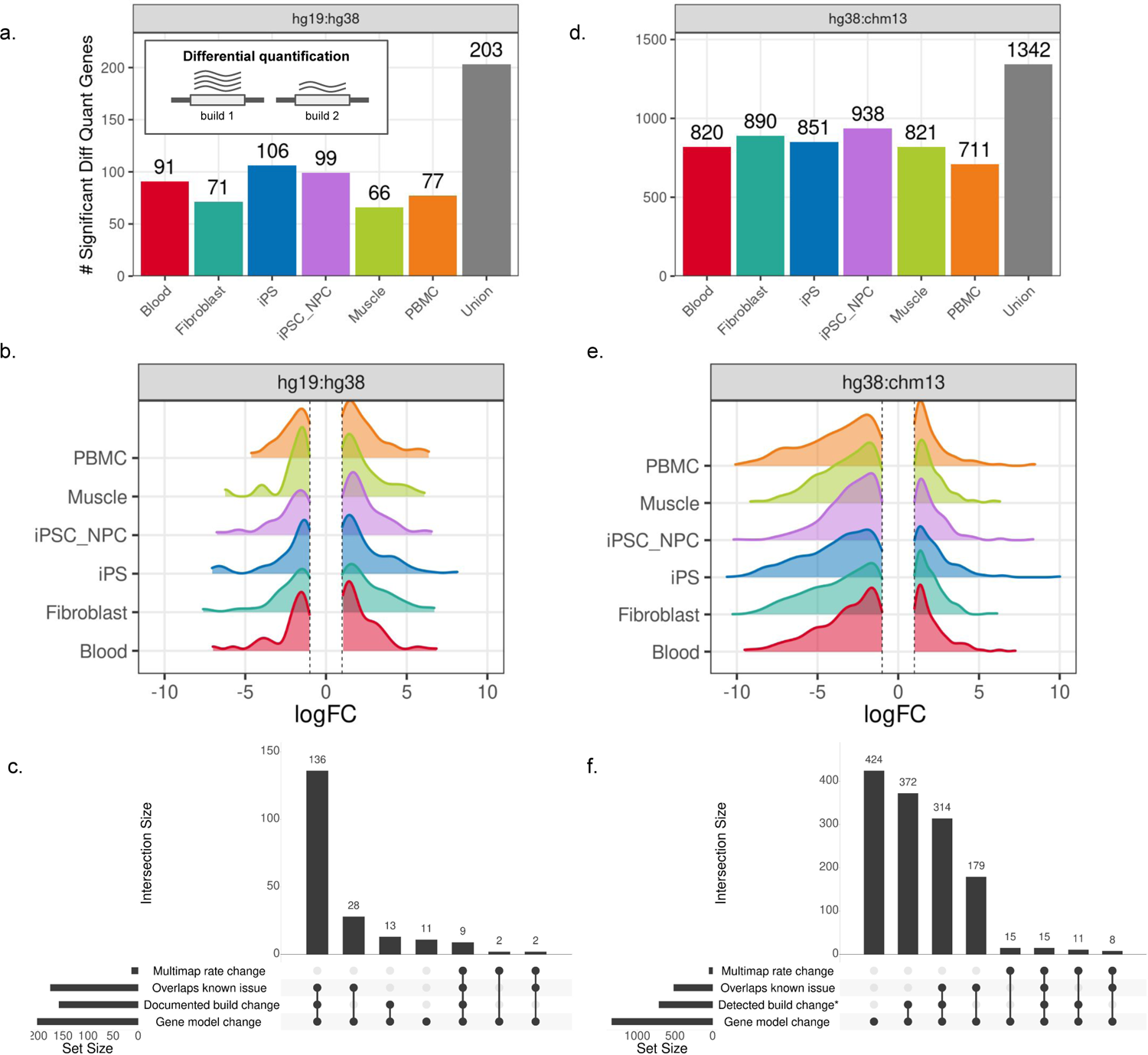
Hundreds of genes are significantly and substantially differentially quantified between builds **a**, The number of genes that were significantly differentially quantified by build (adjusted p-value <0.05 and abs(logFC)>1) between hg19 and hg38 and **d**, between hg38 and chm13 across tissue types. Gray bars on the far right display the union of differentially quantified genes across all tissues. Inset in **a** provides a visual definition of differentially quantified events in which a gene is annotated and sufficiently quantified in both alignments the expression estimates differ. **b,** Distribution of logFC values for significant genes (adjusted p-value < 0.05) across tissues for hg19 compared to hg38 and **e**, hg38 compared to chm13. **c**, Upset plot displaying the putative reasons underlying differences in gene expression estimates between hg19 and hg38, and **f,** hg38 and chm13.

Comparison between hg38 and CHM13 allowed us to test 30,998 genes, including 72.4% (20,256/27,966) of the known protein-coding and lncRNA genes (**Supplementary Table 4**). We identified 1,341 genes with significant and substantial differential quantification in at least one biospecimen type, including 1,132 protein-coding and lncRNA genes **(Figure 3d,e, Supplementary Figure 6b, 7b**). The majority (1,028) of the differentially quantified genes were identified in more than one biospecimen, with 452 observed in all six (**Supplementary Figure 8b**). Over a third (516) of the hg38:CHM13 differentially quantified genes overlapped a documented issue or difficult-to-sequence region in at least one build (**Figure 3f**). Of the 825 genes residing in putatively non-erroneous regions across both builds, most (768 genes) harbor changes in the gene model or overlap a documented assembly change between hg38 and CHM13 (**Figure 3f**). Thus, about 90% of hg38:CHM13 differentially quantified genes may be explained by a change in the gene model or overlap of a documented error in the hg38 reference genome (**Figure 3f**).

Across both build comparisons, 341 differentially quantified genes are implicated in a rare disease with a known molecular basis in the OMIM database (7 hg19:hg38, 262 hg38:CHM13). Additionally, 38 are confidently linked to a disease in the OpenTargets Platform with a score greater than 0.8 (1 hg19:hg38, 38 CHM13:hg38) (**Supplementary Table 1**). We detected a number of genes linked to cancer in the COSMIC database that are significantly and substantially quantified differently by build including 1 in hg19:hg38 and 65 in hg38:chm13^44^. The sole gene in hg19:hg38 is U2AF1, which can cause myelodysplastic syndromes and has been shown to respond to therapeutic targets^45, 46^. New contigs for this gene were added in hg38 build, and as a result there was 7.8x larger expression in hg38 relative to hg19. However due to a number of persisting issues in the hg38 assembly of this region, including false duplications that caused high levels of multimapping, U2AF1 was quantified 103x higher in chm13 than hg38. Additional cancer genes with substantial differential quantification between hg38 and chm13 included EGFR, RB1, KRAS, and BRIP1.

### Multiple disease-relevant genes display build-exclusive expression

Differential quantification can only assess the impact of build choice for genes that are annotated and sufficiently expressed in both builds; therefore we further explored genes excluded from the differential quantification analysis due to insufficient expression levels in just one build per comparison despite presence in both annotations, hereafter referred to as genes with build-exclusive expression (**Figure 4b, Supplementary Table 5**).

**Figure 4.**
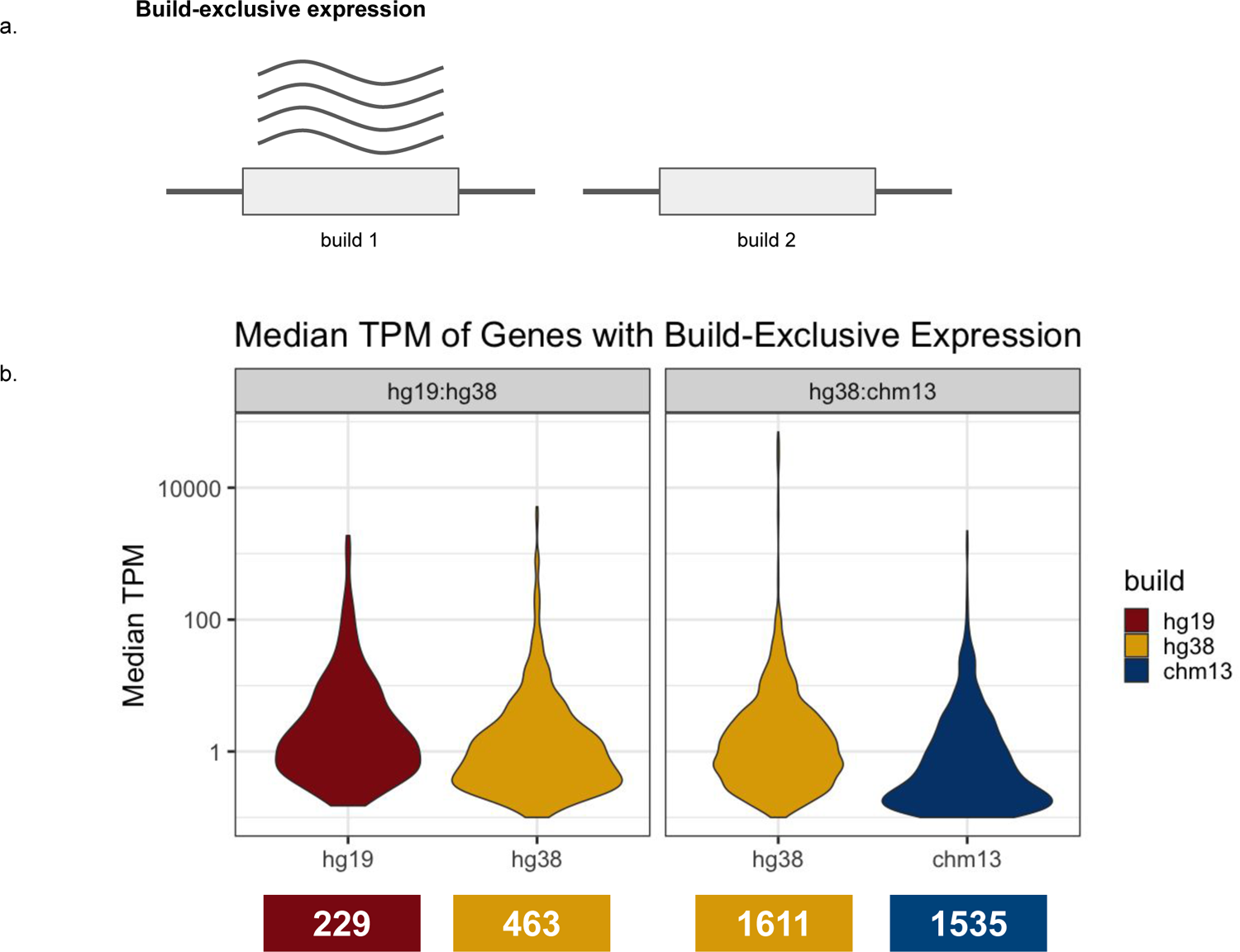
Hundreds of mutually annotated genes show substantial build-exclusive expression **a**, Depiction of build-exclusive expression. **b**, Distribution of median TPM levels of build-exclusive genes on a log scale for the hg19:hg38 comparison (left) and hg38:chm13 comparison (right). The number of build-exclusive genes detected are labelled underneath.

We further explored build-exclusive genes that were not due to a thresholding effect (see **Methods**). When comparing hg19:hg38 we detected 83 genes quantified only in hg19 and 124 only in hg38, 70% and 85% of which overlapped erroneous or blacklisted regions respectively(**Figure 4b**, **Supplementary Figure 9, 10b; Supplementary Table 5**). Within the hg38:CHM13 comparison, 585 mutually annotated genes were only detected from the hg38 alignment and 272 from the CHM13 alignment, 65% and 40% of which were in known erroneous or blacklisted regions respectively (**Figure 4b; Supplementary Figure 9, 10d; Supplementary Table 5**).

The largest reason for build-exclusive expression was due to changes in the gene model between builds, which can impact mappability when aligning to the transcriptome, leading to build-exclusive expression (**Supplementary Figure 10**). The protein-coding gene PDGFRB, is implicated in multiple rare disorders, including Kosaki overgrowth syndrome^43^. Approximately 70% (194/275) of the samples with quantification of this gene in hg38 do not meet the expression threshold in CHM13 in blood. Notably the CHM13v2 annotation for PDGFRB was more complex than the hg38 and hg19 gene models, with three additional transcripts, resulting in higher multimapping rates and making accurate quantification more difficult when performing transcriptome-based alignment.

Known issues and blacklisted regions can also contribute to build-exclusive expression (**Supplementary Figure 10**). BMS1P8, a gene that has been linked to lower survival rates in hepatocellular carcinoma based on hg19-derived expression levels, was only detected in hg19 in fibroblast^47^. However, this association may be an artifact driven by an error in this region in the hg19 reference. While BMS1P8 was annotated in both hg38 and CHM13, the contigs used to construct the region were updated in the hg38 assembly to resolve this hg19 error^43^.

### Transcriptomic outlier detection between genome builds

Transcriptome outliers provide evidence to enable gene prioritization in Mendelian diagnoses^1^. To assess if genome build impacted our ability to detect outliers, we called expression outlier events in blood and fibroblast samples. For each biospecimen we calculated z-scores for each gene-individual pair and defined an outlier as an absolute z-score greater than 3 (see **Methods**); this identified thousands of expression and splicing outlier events in each build (**Table S3**). Of note, splicing outlier detection was reference-free^48^, so discrepancies in splicing outliers between builds were fully attributable to build-based alignment differences.

We assessed the consistency of expression and splicing outlier status (eOutliers and sOutliers) by determining the proportion of outliers in one build that were also outliers in another build (see **Methods**). There were a similar number of outliers between builds (**Figure 5a**). The vast majority of eOutliers were consistent between builds (**Figure 5b, Supplementary Table 6**), and most of the inconsistent eOutliers were the product of a thresholding effect wherein a gene’s z-score in the non-outlier build fell just below the outlier definition cutoff (see **Methods**). In line with our differential quantification results, outliers were more consistent between hg19 and hg38 compared to hg38 and CHM13 (**Supplementary Figure 11**). sOutlier detection was also largely consistent between builds, but was generally less impacted by thresholding effects (**Supplementary Figure 11**). However, some individuals did have dramatic changes in outlier status for a handful genes; genes with large discrepancies (absolute z-score>3 in one build and <1 in the other) accounted for 3-23% of inconsistent eOutliers, and 36-74% of inconsistent sOutliers. We further detected 68 high-confident OMIM eOutlier genes and 99 OMIM sOutlier genes that have at least one outlier substantially different between builds, indicating that consideration of these disease-relevant genes for potential diagnoses may be impacted by build choice (**Supplementary Table 7**). Across both expression and splicing we also detected hundreds of unique genes that were only annotated in hg19, hg38, or CHM13 that were considered outliers, of which a large proportion were labeled as erroneous (**Supplementary Table 6**).

**Figure 5.**
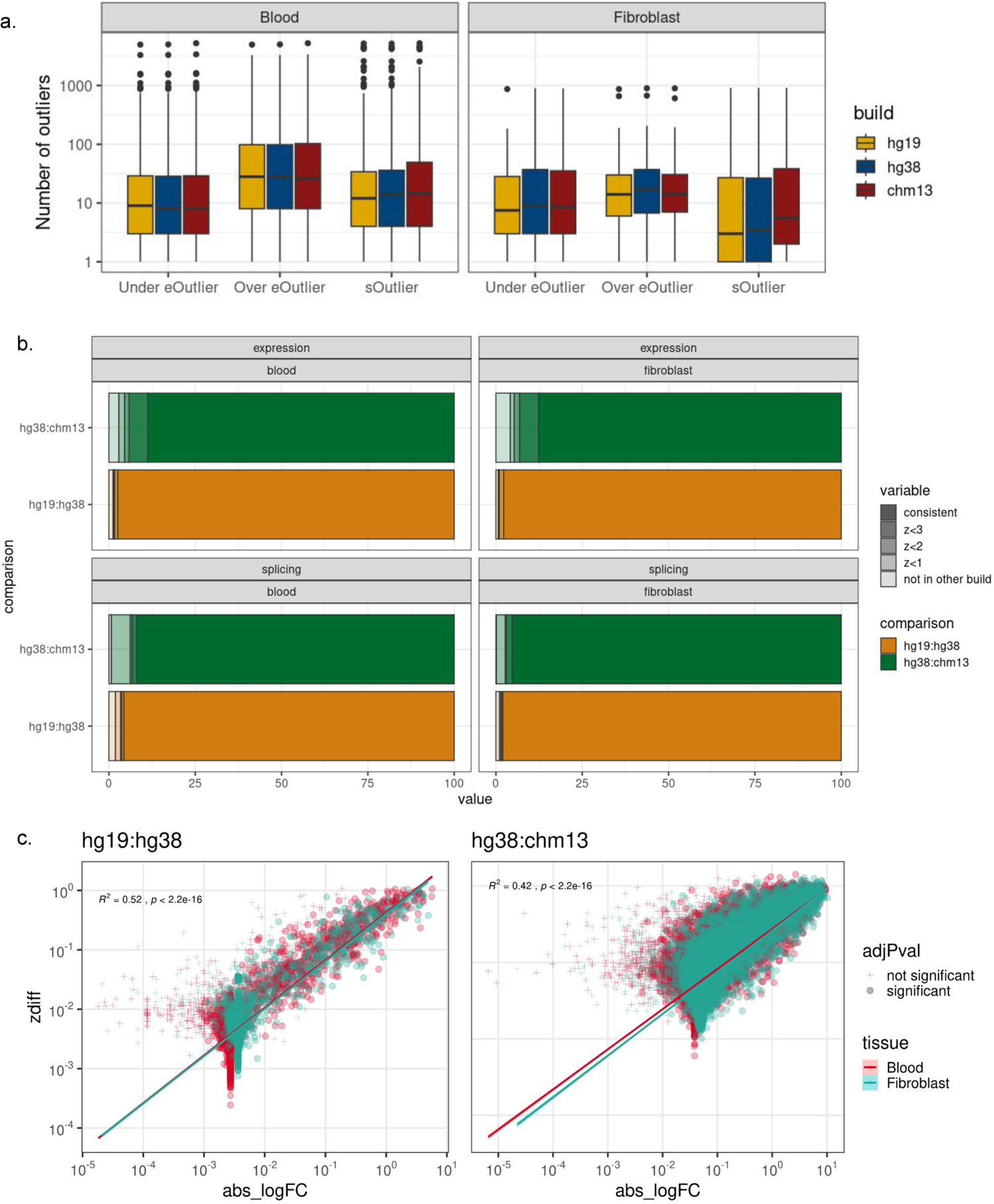
Impact of build selection on expression and splicing outlier detection **a,** Boxplots displaying the number of over-expression, under-expression, and splicing outlier genes per sample detected from data aligned to hg19 (red), hg38 (yellow), and CHM13 (blue). **b**, Expression outlier consistency between hg19:hg38 (left) and hg38:chm13 (right). In orange the outliers that are consistent between hg19 and hg38, and in dark green the number of outliers consistent between hg38 and chm13. In lighter shades, the number of outliers with a z-score greater than 3 in chm13 but less than 3 in hg38 (or greater than 3 in hg38 but less than 3 in hg19) and so forth. The lightest shares are outlier in the reference build (ex chm13) but are NA in the comparison build (ex hg38) due to lack of quantification in that build. This is faceted by tissue type, and expression vs splicing outliers. **c**, Comparison between differential quantification fold change and average absolute z-score change. Each data point is a gene, the x-axis represents the absolute log fold change in the differential quantification results, and the y-axis dictates the average change in z-score between builds for that gene. This is plotted for hg19:hg38 (left) and hg38:chm13 (right), the shape of the point is determined by significance level, and the color of the point is determined by tissue.

We then investigated if there was a relationship between the differential quantification of a given gene (absolute logFC) and its average change in outlier z-score (mean z-difference) between builds. We observed that the degree to which gene quantification differed between builds correlated with larger z-score changes (R^2^ 0.42-0.52, **Figure 5c**), indicating that the more differentially quantified a gene is, the more likely it will be to impact outlier status. Further, genes with large eOutlier changes (absolute z-score >3 and <1 in complementary builds in at least one sample) were more likely to be differentially quantified genes (average logFC 1.12 hg19:hg38 and 1.80 hg38:CHM13 in large eOutlier change genes, 0.02 hg19:hg38 and 0.18 hg38:CHM13 for genes without large eOutlier changes; **Supplementary Figure 12**).

### Impact of genome build on interpretation of clinically relevant genes

Clinicians may often only have time to systematically evaluate a handful of candidate genes. While there are many ways to prioritize this top candidate list, one approach is to identify genes with the greatest aberrant expression events based on expression or splicing outlier scores. Given the time and resources required for manual curation, it is important to consider that ranking a gene as the 18th largest outlier compared to the 32nd will impact its potential to be reviewed. While we noted some genes changed in their outlier status between builds, here we assessed how the rank of genes prioritized for manual curation changed between builds.

We found that transcriptome-guided candidate gene lists in cases, which we defined as the top 20 most extreme expression and splicing outlier genes per case, were largely consistent between builds, though more so for hg19:hg38 (**Figure 6a,c**). For genes that were in the top 20 list in one build, but not in the top 20 list in the other build, the ranks were still near the threshold in hg19:hg38, but changed more so in hg38:CHM13 (**Figure 6b,d**).

**Figure 6.**
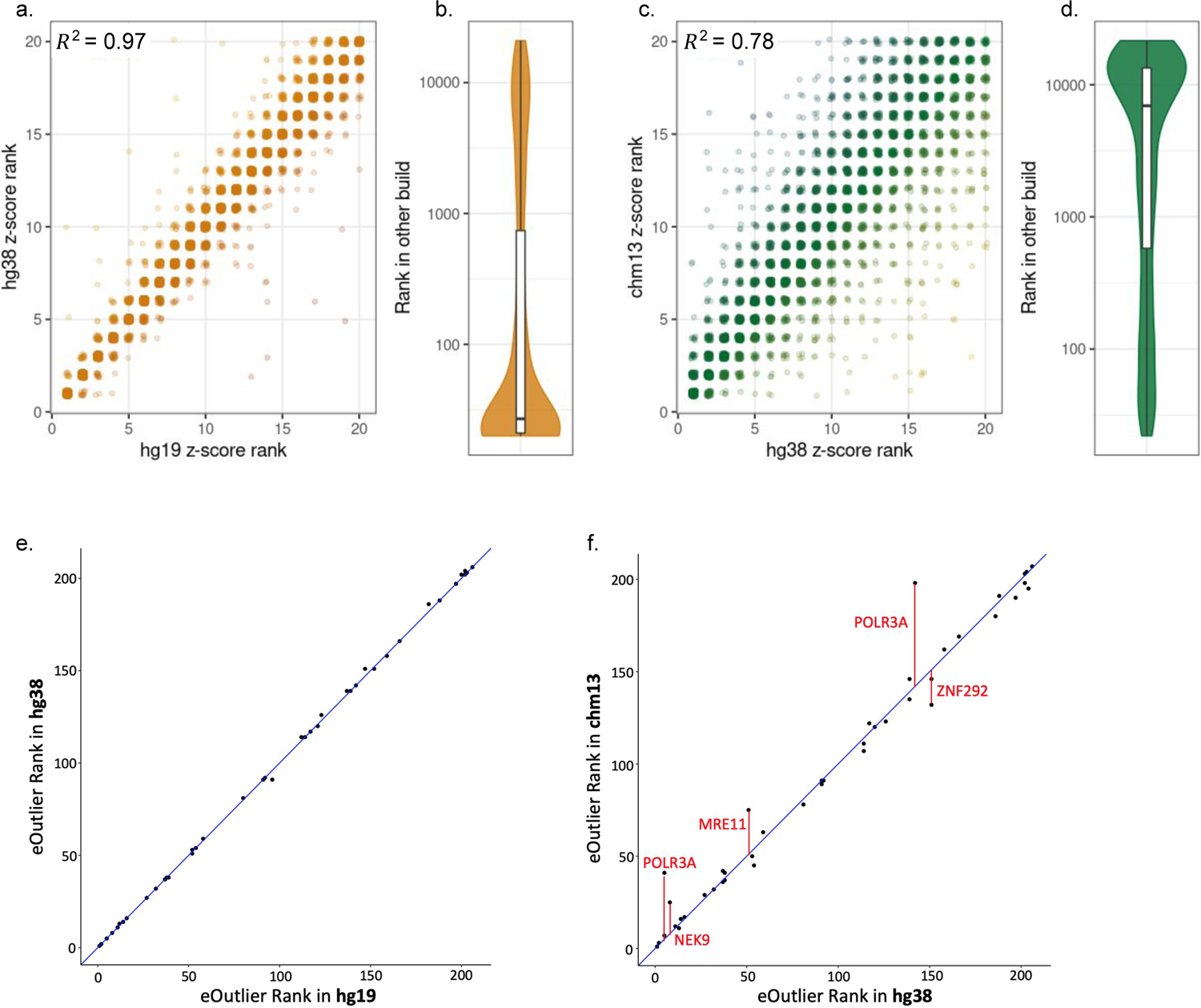
Build selection impacts transcriptome-guided gene prioritization **a**, Comparison of the ranked z-scores for genes in the top-20 expression outlier lists from both the hg19 and hg38 alignments across all cases (Pearson correlation *R*^2^ = 0.97). **b,** The distribution of z-score ranks for genes that were only in a case’s top-20 list in one build for the hg19:hg38 comparison. **c**,Ranked z-scores for genes in the top-20 expression outlier lists from both the hg38 and chm13 alignments across all cases Pearson correlation for z-score ranks in both top-20 lists *R*^2^ = 0.78). **d**, The distribution of z-score ranks for genes that were only in a case’s top-20 list in one build for the hg38:chm13 comparison. **e**, Diagnostic gene outlier ranks among the top 250 phenotype-prioritized genes across 44 samples from 36 cases with underexpression based on hg19 alignment (x-axis) and hg38 alignment (y-axis). **f**, Diagnostic gene outlier ranks among the top 250 phenotype-prioritized genes across 44 samples from 36 cases with underexpression based on hg38 alignment (x-axis) and chm13 alignment (y-axis). The top five gene-sample pairs with the most extreme residuals are highlighted.

Although outlier ranks were largely consistent for genes annotated in both builds, candidate eOutlier gene lists for 92 cases included 83 distinct annotation-specific genes, the majority of which overlapped a known issue or blacklisted region (**Supplementary Table 8**). Similarly, sOutlier candidate gene lists for 173 cases included 132 annotation-specific genes with many also overlapping issue or blacklisted regions (**Supplementary Table 8**). In the hg38-specific gene (relative to CHM13) SIK1, we detected over-expression of this gene in two cases (which had no related phenotype terms to the associated disorder) and two controls, including for one sample in which it was the highest ranked expression outlier, reiterating our previously made case that SIK1 should not be currently evaluated in hg38 (Figure 2d). Many of these annotation-specific outliers were ranked in the top twenty largest outliers for a sample, potentially impacting a transcriptomics-first approach. Therefore, these annotation-specific genes can lead to erroneous candidates.

Finally, we assessed how build selection impacted prioritization of known causal genes in 44 samples across 38 solved cases. In practice, candidate genes are often prioritized first based on phenotypic and genotypic relevance and then transcriptomic data is to provide further functional evidence. We generated a list of 250 genes for each affected proband derived from their unique symptoms using Phen2Gene^49^, which were then ranked based on expression and splicing z-scores across the three genome builds. While outlier ranks among the diagnostic genes were largely consistent between builds (**Figure 6 e,f**), we noticed genome build impacted our ability to prioritize the diagnostic gene for multiple cases. We identified a teenage female with primarily neurological symptoms and disease-causing mutations in POLR3A, a gene associated with multiple nervous system conditions. POLR3A ranked in the top 5 most aberrantly under-expressed genes for this case in both hg19 and hg38 (z-scores −1.6).

However, its expression was within a standard deviation of the mean when aligned to CHM13 (z-score −0.7) and it ranked 41st across all genes tested for this individual. POLR3A showed significantly higher expression in hg38 compared to CHM13 (absolute logFC = 1.8), which was likely the result of higher rates of multi-mapping in the region in the CHM13 alignment (36% multi-mapped hg38, 88% multi-mapped CHM13) due to a difference in the number of transcripts, consistent with the region’s classification as a blacklisted “High Signal Region” in CHM13. In a case with a known causal gene, we also identified a splicing outlier (z=2) in the non-causal gene NOTCH2 in hg19 only, in which it was the top ranked outlier in hg19 despite showing no evidence of aberrant splicing in hg38 or CHM13. NOTCH2 overlaps a region that included a bacterial contaminant sequence in hg19 that was corrected in hg38 and may account for the erroneous splicing signal. Thus, we found that build selection impacted both our ability to accurately prioritize the true causal gene and eliminate false positives.

## Discussion

Transcriptome sequencing is a widely-used assay that complements the study of genome function, disease mechanisms and diagnosis. It has been increasingly used to supplement exome-negative rare disease cases, with a diagnostic yield between 7-36% depending on the cohort^1^. As an increasingly important clinical tool in rare disease diagnosis, it is important to understand the robustness of transcriptomic data with respect to genome build. The human reference genome has undergone numerous releases since its debut in 2001, with the most recent release of CHM13v2.0 in 2022^28^. Yet clinical uptake of new reference genomes has not kept pace; transitioning infrastructure to new genome builds is expensive and re-interpreting results is time-consuming^11^. Therefore, we provide an annotated resource of build-affected genes to aid in the design and interpretation of reference-aligned RNA-seq (**Supplementary Table 1**).

We describe three primary ways in which genes differ by build: annotation-specific quantification in which a gene is only annotated in one of two builds, build-dependent quantification in which a mutually annotated gene has significantly different expression estimates, and build-exclusive quantification in which a mutually annotated gene fails to meet the expression threshold in one of two builds. Across six routinely-collected biospecimens and three builds, we identified 2,800 genes with build-dependent quantification including 1,391 protein coding genes, and 341 known Mendelian disease genes. We were well-powered to assess patterns of aberrant expression and splicing in two of the most routinely-collected biospecimens, whole blood and patient-derived fibroblasts. Larger sample sizes will be required to assess the impact of genome build on tissue-specific aberrant expression and splicing events from biosamples that are more clinically or experimentally challenging to obtain.

We suggest caution when examining annotation-specific and build-exclusive genes; a large proportion overlap with erroneous or blacklisted regions and are non-protein coding or lncRNA genes. We further identified build-exclusive genes that were likely due to multimapping; when mapping to complicated gene models, we suggest using a quantification method that employs a genome-based alignment to improve accuracy. These annotation-specific and build-exclusive genes can appear in the top candidate gene list for rare disease patients. We observe that the extent of differential quantification of a gene is correlated with the average change in z-score for a given gene. Additionally we found that 92% of known discrepant regions from Li et al. between hg19 and hg38 were either differentially quantified or had large outlier differences^30^. While many genes had consistent outlier detection across genome builds, we recommend careful evaluation of genes in this list.

While this study focused on the use of transcriptomics for Mendelian disease diagnostics and discovery, these analyses have implications for any human genetics study using RNA-seq^50^. In clinical settings RNA-seq is used to clarify variant interpretation for a range of common hereditary disorders; Invitae estimated RNA-seq would result in reclassification of a splicing variant of unknown significance in 1.7% of individuals in their database^51^. Similar to the rare disease space, cancer diagnostics is increasingly using RNA-seq as a complementary approach to DNA sequencing^52^. One study estimated additional RNA-seq information could impact clinical management for 1 in 43 cancer patients and family members^53^. Our study identified 68 cancer-related genes with build-dependent expression estimates, emphasizing the importance of genome build selection in accurate assessment of transcriptomic biomarkers. These impacts can extend beyond cancer to any area of transcriptome analysis, therapeutic, and diagnostic use.

Throughout, we have demonstrated that hg19 and hg38 RNA-seq results were more congruous than hg38 and CHM13, consistent with the greater similarity in how the references were constructed. While hg38 included an additional 75 Mb of genomic sequence not present in hg19, the CHM13v2 assembly added nearly an additional 200 Mb^43, 54^. Additionally, CHM13v2 is composed of data from a single female of primarily European ancestry^55^, while hg19 and hg38 are primarily based on the genome of one male of admixed African and European ancestry but include additional information from diverse samples from the 1000 genomes project to fill in gaps and fix erroneous regions^43^. Importantly, this means CHM13v2 results might be less reliable for non-European ancestries. Efforts such as the first draft of the pangenome project are expected to improve sequencing alignment for individuals with underrepresented ancestries^56, 57^.

Ultimately, we provide a resource to empower researchers to make the best decisions for their datasets and cases. This list of genes can then be more carefully quantified manually or with tools such as FixItFelix, which can remap erroneous regions to enable better results^58^. We ultimately recommend using our resource to flag genes of interest that might be impacted by build or to choose the best build for a given project.

## Material and methods

### Study cohort

We generated paired-end RNA-seq data on 386 distinct samples from 316 rare disease patients and family members from the Undiagnosed Diseases Network (UDN). Sequenced biospecimens include whole blood (n=283), fibroblasts (n=66), peripheral blood mononuclear cells (PBMCs, n=12), muscle (n=11), induced pluripotent stem cell lines (iPSC, n=8), and iPSC-derived neural progenitor cells (iPSC_NPC, n=6). Samples were ascertained from 316 individuals including 204 rare and undiagnosed cases affected by primarily neurological, musculoskeletal, or immune-related conditions (Figure 1a). Of these samples, 243 are novel and 143 were previously published in Fresard et al^15^; 35 of the individuals in this study are also enrolled in the Genomics Research to d the Genetics of Rare diseases (GREGoR) Consortium. Ethical and research approval was obtained from the National Human Genome Research Institute (NHGRI) Institutional Review Board (IRB) under Protocol 15-HG-0130 and Stanford University IRB (Protocols 23066, 32641, 38046, and 60837). Informed consent was obtained from all participants.

### Transcriptome sequencing

#### Sample collection

In total, RNA-sequencing was performed on 270 samples from 204 affected individuals, and 116 samples from 112 unaffected family members. This included 14 cases from the Utah UDN site in which the Paxgene samples and fibroblast cell lines were collected and established at Utah, then the processed RNA was shipped to Stanford. Whole blood samples were collected and processed in Paxgene RNA tubes at Stanford. Globin mRNA was removed from whole blood samples using NUGEN (n=92) or GLOBINClear for the remainder (n=191).

#### RNA-sequencing library preparation and sequencing

Two protocols were used for library preparation and sequencing given a switch to increased automation. cDNA libraries for the first 258 samples were generated using the Illumina TrueSeq Stranded mRNA Sample Prep Kit protocol, and dual indexed. Bioanalyzer and Qubit were used to determine proper library dilution and balance samples across sequencing runs. Samples were pooled and sequenced on an Illumina NextSeq 500 across 14 distinct runs including between 15 and 20 samples. Two runs generated 75-bp paired-end reads, and the other twelve generated 150-bp paired-end reads.

An additional 128 samples were processed with the Universal Plus™ mRNA-Seq with NuQuant library prep protocol from Tecan, which includes globin and ribosomal RNA depletion following the same protocol as Amar et al^59^. These samples were processed in an automated protocol on a Biomek i7 robotic liquid handling system. Library quality was evaluated based on fragment analyzer tracings, and cDNA concentration was determined with Qubit. Libraries were normalized by molarity prior to sequencing on a Illumina Novaseq 6000 in two runs containing 97 and 34 UDN samples respectively.

### Pipeline

#### Overview

We demultiplex BCL data into FASTQ files, align to the genome, quantify the reads, and call splicing and expression outliers (**Supplementary Figure 2**). These steps are detailed in the following sections.

#### Transcriptome QC and Alignment

Genome build reference files were downloaded from GENCODE^60^. We used the primary assembly for hg19 and hg38 (), and CHM13v2 with the Y-chromosome masked. We used GENCODEv35lift37 and GENCODEv35 primary genome annotations (chromosomal regions) for hg19 and hg38, respectively, and the GENCODEv35 CAT/Liftoffv2.0 annotation for CHM13. The full download links are available in the study github (https://github.com/raungar/build_rnaseq_paper_public).

The gff3 file for hg19 was generated from the gtf file using gffread^61^. RSEM references were prepared with rsem/1.3.1^62^. An annotation file for STAR/2.8.4a^33^ was created for an overhang of 99 for files with a read length greater than 100 and an overhang of 75 for those with a read length of 76.

FASTQ files generated from demultiplexing the raw BCL data using bcl2fastq^63^ and were trimmed using cutadapt^64^ with a minimum trim length of 20 for reads less than 100bp and 50 for those greater than or equal to 100bp. Reads were aligned with STAR (see github for full parameters). Optical duplicates were filtered from aligned bam files using Picard^65^ and a pixel distance of 12000px for samples sequenced on the Novaseq machine (n=128 samples), and 100px for samples sequenced on the NextSeq500 (n=251).

#### Quantification

To assess build-dependent quantification differences of uniquely mapped reads, samtools^66^ was used to filter uniquely-mapped reads only. Filtered bam files were then prepared for and processed by RSEM^62^ to obtain read counts per transcript and per coordinate site. As only uniquely mapped reads were included, the estimated counts provided by RSEM are equivalent to raw counts.

#### Expression Outlier Calling

We adapted the outlier calling method from Frésard et al^15^. We called outliers for each biospecimen with at least 30 samples and required a minimum of three samples per batch. For each gene, at least 20% of samples must have a TPM of 0.15 or larger. If there are fewer than 150 samples, then at least 30 samples must have a TPM of 0.15 or larger for a given gene to ensure an ability to perform outlier calling. The counts are then log transformed, genes with zero variance are removed, and then each gene is scaled and centered. Batch, RIN, and sex are regressed out and the residuals are again normalized. Given the variance has been adjusted to be 1, we expect each batch to also have a variance of 1. If a given batch has a low variance (less than 0.1) for a given gene, the samples in the batch are removed as it is likely an artifact. The z-score for this gene is recalculated and the low-variance samples are set to NA.

#### Splicing Outlier Calling

Splice junctions were extracted and annotated using regtools^67^. Introns were clustered using LeafCutter^48^ and outliers were called using LeafCutterMD^68^. LeafCutterMD detects how likely the reads overlapping a cluster of exon-exon junctions for a given sample comes from the same distribution as all other samples. Therefore, a p-value is reported for each exon-exon junction per sample. We adjusted the p-value for multiple-testing using the Benjamini-Hochberg method, and then convert it to a z-score. Finally, to report splicing outliers at a gene-level we annotated genes for each exon-exon junction using bedtools^69^ and reported the maximum z-score that overlaps with this gene.

### Comparison of genome annotations and gene models between builds

To investigate the relationship between the hg38 gene models and the hg19 and CHM13 gene models, we compared the GENCODEv35 primary annotation for hg38 and corresponding annotations for hg19 (GENCODEv35lift37)^70^ and CHM13 (T2T consortium CAT-LIFTOFF)^71^. We chose GENCODEv35 as it was used to construct the CHM13 annotation model, and therefore would minimize differences due to annotation to focus on the build comparison. To identify mutually annotated genes, we leveraged the GENCODE remapping results for the the GENCODEv35 liftover to hg19^70^, and the remapping metrics provided in the CHM13v2 CAT-LIFTOFF annotation file. For hg38:CHM13, gene models were considered common if the hg38 gene was used as the source gene for the CHM13 gene models (as indicated in the CHM13 GTF file); for expanded pseudogene families, the parent genes annotated in hg38 and CHM13 were considered common, and the additional pseudogenes added in CHM13 were considered annotation-specific; genes with no corresponding gene model in the other build were also considered annotation-specific. Several methods were employed to establish relationships between hg38 and hg19 gene models, to account for the automated and manual annotation procedures employed during the liftover process. By default, gene models were assigned the relationship indicated in the liftover results file; if no relationship was indicated, the relationship was established manually based on the consistency of gene id, gene version, and gene symbol. Genes where no relationship could be established were considered specific to one or the other annotation.

We characterized each gene in terms of structure and sequence by counting the number of transcripts and exons within each gene model and calculating their lengths using positions indicated in the respective GTF files. Transcript length was considered as the sum of the size of its constitutive exons, and we extracted the sequence of each exon using bedtools v2.25.0 toolset and considering the orientation of the gene model (function: getfasta -s)^69^. We compared the gene models for genes present in multiple annotations in terms of structure conservation by calculating the difference in (a) the number of transcripts, (b) the number of exons per transcript, and (c) the difference in length between exons. Gene model comparison results are summarized in **Supplementary Figure 3**, and included per-gene in **Supplementary Table 1**.

For mutually annotated genes with the same structure, we calculated sequence variation by using the Jaro-Winkler distance (R-package RecordLinkage v0.4.12.4)^35, 72^, which was performed at the level of the constitutive exon, transcript (average of these exons), and gene level (average of these transcripts). The Jaro-Winkler scores are included in **Supplementary Table 1** for genes with identical gene models and a similarity score below 1..

### Differential quantification

We performed paired differential expression analysis for hg19 vs hg38 (hg19:hg38) and hg38 vs CHM13 (hg38:CHM13) across all samples within each of the six biosample types. The sample sizes for each analysis are presented in **Supplementary Table 2**. We filtered to genes with a single gene version present in each build annotation, and for which at least 30% of samples had at least 0.1 counts per million in each build. Y-chromosome genes were excluded from the hg38:CHM13 comparison due to the incomplete Y-chromosome annotation for CHM13 at the time this analysis was performed.

Paired differential expression analysis was performed using the LIMMA-DREAM framework, which allows for a generalized linear model with repeated measures^41, 42^. Since the same RNA-seq FASTQs were aligned to each of the three genome builds, there is no biological difference between the alignments, and the genome build can be considered a computationally-produced condition in a repeated measures study design with identical underlying samples. This was modeled using a linear mixed model, in which build was treated as a fixed effect, sample was treated as a random effect, and expression (TPM) was the dependent variable^41, 42^.

### Identification of build-exclusive events

We identified genes that were either annotation-specific, or build-unique. Annotation-specific genes were those that did not exist in at least one build’s annotation but were present in another. Similarly genes were considered to be build-unique if there was not sufficient quantification (expression) in at least one build, while there was sufficient quantification in another. A gene is considered expressed in a given build if 30% of individuals express this gene at a TPM greater than 0.1. We defined build-exclusive genes to be those in which a gene was quantified in one build but not another. We defined a build-exclusive gene to be caused by a threshold effect if the number of individuals with a TPM greater than 0.1 was within two people of the minimum needed individuals.

### Defining genes of medical interest

Multiple databases characterizing gene-phenotype associations were queried to identify medically relevant genes. The Online Mendelian Inheritance in Man (OMIM) database was utilized to identify genes linked to known rare and Mendelian disorders; rare disease genes were defined as those with an OMIM gene-phenotype relationship score of 3 or 4, indicating that the molecular basis of the disease is known or is caused by chromosomal deletion or duplication^73^. Cancer-related genes were identified from the Catalog of Somatic Mutations in Cancer (COSMIC) data^74^, and additional disease-related genes were identified by querying the OpenTargets platform^75^ (filtering to genes with a phenotype evidence score of at least 0.8 out of a maximum score of 1), and the ClinVar database^76^. A gene was considered a known disease gene if it was linked to a disease phenotype in any of these sources.

### Identification of genes overlapping regions with known issues

Issue-prone regions of the genome for each build were defined based on both official issue reports from the consortium that produced the assembly (hg19 and hg38: Genome Reference Consortium^9, 43^; CHM13: Telomere2Telomere Consortium^28^), and region blacklists generated by independent sources^77, 78^. The hg19 and hg38 exclusion regions (previously “blacklisted regions”) were defined by ENCODE as difficult-to-sequence regions with tendencies towards high multimapping rates or high mapping variability^77^. Bed files delineating the ENCODE blacklisted regions for hg19 and hg38 were accessed for this study on January 23, 2023. As no official ENCODE exclusion list for CHM13 was available at the time of this publication, the corresponding blacklisted regions for CHM13 were obtained as a bed file on February 21, 2023 from excluderanges, a bioconductor package for tracking problematic genomic regions across genome assemblies^78, 79^.

The genomic regions with known issues in hg19 and hg38, as defined by the Genome Reference Consortium, were downloaded from the UCSC Genome Browser GRC incident tracks on January 25, 2023^80^. BigBed browser track files were converted to bed files with bigBedToBed from the UCSC Genome Browser tools^81^, and used to annotate the appropriate genome annotation GTF files using bedtools intersect^69^. We defined a gene as overlapping a known issue in hg19 or hg38 if it intersected with any issue type and the status for the issue was not marked “resolved”, or if the gene overlapped an ENCODE blacklisted region.

Problematic regions that have been identified in the CHM13 assembly were downloaded as bed files from CHM13 issues github repository on January 25, 2023^82^. Bedtools intersect with the CHM13v2.0 annotation GTF file was used to link genes and problematic regions^69^. A gene was considered to fall within a CHM13 issue-prone region if it overlapped with either one of the exclusion regions from excluderanges or one of the regions reported to harbor a known issue^78, 79^.

### Characterizing genes impacted by changes in genome assembly

To identify regions of the reference genome that were updated between hg19 and hg38, the UCSC Genome Browser tracks delineating regions where the reference sequence construction differed (hg38ContigDiff.txt.gz and hg19ContigDiff.txt.gz) were downloaded on August 1, 2022^83^. The text files provided a set of impacted genomic ranges in hg38 and hg19 coordinates, respectively, and a score of 0, 500, or 1000 which was recoded in accordance with the UCSC table schema for the track^84^. A score of 0 indicates that a new contig was added in the hg38 construction of the region to update the sequence or address gaps present in the hg19 assembly. A score of 500 corresponds to regions where different portions of the same contig was used to construct the same region, potentially leading to differences in sequence. Finally, a score of 1000 indicates that hg19 sequence errors have been corrected with updated contigs.

Because hg38 and CHM13 were not constructed from the same collection of BAC clone contigs, an equivalent table of contig usage was not available. Instead, we leveraged the hg38-CHM13 alignment tracks provided by UCSC as a bed file, which was downloaded on October 19, 2022 and used to define regions present in the CHM13 assembly but absent in hg38 (a.k.a. “non-syntenic” regions)^83–85^. Additionally, information about CHM13-specific reference artifacts and variations were included for 4,964 medically-relevant genes, as summarized in supplementary table 13 provided by Aganezov et al (2022)^54^.

### Outlier comparison

We sought to identify when an outlier is called in a build-specific way. For outliers in a given build, we identified the z-score in the comparison build. We evaluated this bidirectionally; for example between hg19:hg38 of the hg19 outliers we examine the z-score in hg38, and of the hg38 outliers we examine the z-scores in hg19.

We next identified which genes have large outlier status between builds were differences which were not due to thresholding. Given our threshold for outliers status is a z-score of 3, a gene might have a z-score of 2.9 in hg19 but a z-score of 3.1 in hg38 and would be considered an outlier in a specific build despite it not being biologically meaningful. Therefore, we only considered large outlier changes between builds, defined such that the absolute z-score must be greater than 3 in one build, and less than 1 in the other, hereafter referred to as inconsistent outliers.

## Data and code availability

Our pipeline is fully available at https://github.com/raungar/build_rnaseq_paper_public. Data is currently available or being uploaded for the UDN (phs001232.v5.p2) and GREGoR (phs003047.v1.p1) on dbGaP.

## Acknowledgments

We appreciate members of the Montgomery Lab, Stanford Genetics Department, and GREGoR consortium who have provided valuable feedback throughout this development of this project. This work utilized computing resources provided by the Stanford Genetics Bioinformatics Service Center, supported by NIH Instrumentation Grant S10 OD025082, and would not have been possible without the support of the Stanford SCG cluster system administrators. We additionally would like to thank Shruti Marwaha, Chloe Reuter, Jennefer Carter, and Gyu Kim. We would like to acknowledge and thank the Utah UDN site, including Lorenzo Botto, Ashley Andrews, Erin Baldwin, for providing several samples included in this study. Several figures were made with BioRender. Research reported in this manuscript was in part supported through the Undiagnosed Diseases Network by the NIH Common Fund through the Office of Strategic Coordination, Office of the NIH Director, and the National Institute Of Neurological Disorders And Stroke under Award Numbers U01HG010217 and U01HG010218 This publication was supported in part, by the National Human Genome Research Institute of the National Institutes of Health through the following grants, as part of GREGoR Consortium: U01HG011762. R.A.U., P.C.G, T.D.J were further funded by the Stanford Genome Training Project (T32HG000044). The content is solely the responsibility of the author and does not necessarily represent the official views of the National Institutes of Health.

## Author Contributions

R.A.U., P.C.G., and S.B.M. conceived the study. R.A.U., P.C.G, T.D.J., F.D., and S.B.M. significantly contributed to study design, with feedback from D.E.B, J.A.B, and M.T.W. improving the analyses and focus throughout. Pipelines were developed by R.A.U. and P.C.G. with contributions from T.D.J. Analyses and figures for these analyses were generated by R.A.U., P.C.G, T.D.J., and F.D. Patients were seen by J.A.B., M.T.J, and D.E.B. and samples were processed by K.S.S. and C.A.J. The manuscript was primarily written and figures generated by R.A.U. and P.C.G, with major feedback also provided by S.B.M. All authors provided feedback on the manuscript to improve it.

## Undiagnosed Diseases Network Banner Authorship

Maria T. Acosta, David R. Adams, Ben Afzali, Ali Al-Beshri, Aimee Allworth, Raquel L. Alvarez, Justin Alvey, Ashley Andrews, Euan A. Ashley, Carlos A. Bacino, Guney Bademci, Ashok Balasubramanyam, Dustin Baldridge, Jim Bale, Michael Bamshad, Deborah Barbouth, Pinar Bayrak-Toydemir, Anita Beck, Alan H. Beggs, Edward Behrens, Gill Bejerano, Hugo J. Bellen, Jimmy Bennett, Jonathan A. Bernstein, Gerard T. Berry, Anna Bican, Stephanie Bivona, Elizabeth Blue, John Bohnsack, Devon Bonner, Nicholas Borja, Lorenzo Botto, Lauren C. Briere, Elizabeth A. Burke, Lindsay C. Burrage, Manish J. Butte, Peter Byers, William E. Byrd, Kaitlin Callaway, John Carey, George Carvalho, Thomas Cassini, Sirisak Chanprasert, Hsiao-Tuan Chao, Ivan Chinn, Gary D. Clark, Terra R. Coakley, Laurel A. Cobban, Joy D. Cogan, Matthew Coggins, F. Sessions Cole, Brian Corner, Rosario I. Corona, William J. Craigen, Andrew B. Crouse, Vishnu Cuddapah, Michael Cunningham, Precilla D’Souza, Hongzheng Dai, Surendra Dasari, Joie Davis, Margaret Delgado, Esteban C. Dell’Angelica, Katrina Dipple, Daniel Doherty, Naghmeh Dorrani, Jessica Douglas, Emilie D. Douine, Dawn Earl, Lisa T. Emrick, Christine M. Eng, Kimberly Ezell, Elizabeth L. Fieg, Paul G. Fisher, Brent L. Fogel, Jiayu Fu, William A. Gahl, Rebecca Ganetzky, Emily Glanton, Ian Glass, Page C. Goddard, Joanna M. Gonzalez, Andrea Gropman, Meghan C. Halley, Rizwan Hamid, Neal Hanchard, Kelly Hassey, Nichole Hayes, Frances High, Anne Hing, Fuki M. Hisama, Ingrid A. Holm, Jason Hom, Martha Horike-Pyne, Alden Huang, Yan Huang, Anna Hurst, Wendy Introne, Gail P. Jarvik, Jeffrey Jarvik, Suman Jayadev, Orpa Jean-Marie, Vaidehi Jobanputra, Emerald Kaitryn, Oguz Kanca, Yigit Karasozen, Shamika Ketkar, Dana Kiley, Gonench Kilich, Shilpa N. Kobren, Isaac S. Kohane, Jennefer N. Kohler, Bruce Korf, Susan Korrick, Deborah Krakow, Elijah Kravets, Seema R. Lalani, Christina Lam, Brendan C. Lanpher, Ian R. Lanza, Kumarie Latchman, Kimberly LeBlanc, Brendan H. Lee, Richard A. Lewis, Pengfei Liu, Nicola Longo, Joseph Loscalzo, Richard L. Maas, Ellen F. Macnamara, Calum A. MacRae, Valerie V. Maduro, AudreyStephannie Maghiro, Rachel Mahoney, May Christine V. Malicdan, Rong Mao, Ronit Marom, Gabor Marth, Beth A. Martin, Martin G. Martin, Julian A. Martínez-Agosto, Shruti Marwaha, Allyn McConkie-Rosell, Alexa T. McCray, Matthew Might, Mohamad Mikati, Danny Miller, Ghayda Mirzaa, Eva Morava, Paolo Moretti, Marie Morimoto, John J. Mulvihill, Mariko Nakano-Okuno, Stanley F. Nelson, Serena Neumann, Donna Novacic, Devin Oglesbee, James P. Orengo, Laura Pace, Stephen Pak, J. Carl Pallais, Neil H. Parker, LéShon Peart, Leoyklang Petcharet, John A. Phillips III, Jennifer E. Posey, Lorraine Potocki, Barbara N. Pusey Swerdzewski, Aaron Quinlan, Daniel J. Rader, Ramakrishnan Rajagopalan, Deepak A. Rao, Anna Raper, Wendy Raskind, Adriana Rebelo, Chloe M. Reuter, Lynette Rives, Amy K. Robertson, Lance H. Rodan, Martin Rodriguez, Jill A. Rosenfeld, Elizabeth Rosenthal, Francis Rossignol, Maura Ruzhnikov, Marla Sabaii, Jacinda B. Sampson, Timothy Schedl, Kelly Schoch, Daryl A. Scott, Elaine Seto, Vandana Shashi, Emily Shelkowitz, Sam Sheppeard, Jimann Shin, Edwin K. Silverman, Giorgio Sirugo, Kathy Sisco, Tammi Skelton, Cara Skraban, Carson A. Smith, Kevin S. Smith, Lilianna Solnica-Krezel, Ben Solomon, Rebecca C. Spillmann, Andrew Stergachis, Joan M. Stoler, Kathleen Sullivan, Shirley Sutton, David A. Sweetser, Virginia Sybert, Holly K. Tabor, Queenie K.-G. Tan, Amelia L. M. Tan, Arjun Tarakad, Herman Taylor, Mustafa Tekin, Willa Thorson, Cynthia J. Tifft, Camilo Toro, Alyssa A. Tran, Rachel A. Ungar, Adeline Vanderver, Matt Velinder, Dave Viskochil, Tiphanie P. Vogel, Colleen E. Wahl, Melissa Walker, Nicole M. Walley, Jennifer Wambach, Michael F. Wangler, Patricia A. Ward, Daniel Wegner, Monika Weisz Hubshman, Mark Wener, Tara Wenger, Monte Westerfield, Matthew T. Wheeler, Jordan Whitlock, Lynne A. Wolfe, Heidi Wood, Kim Worley, Shinya Yamamoto, Zhe Zhang, Stephan Zuchner

## References

1. Montgomery, S. B., Bernstein, J. A. & Wheeler, M. T. TOWARDS TRANSCRIPTOMICS AS A PRIMARY TOOL FOR RARE DISEASE INVESTIGATION. Mol. Case Stud. mcs.a006198 (2022) doi:10.1101/mcs.a006198.

2. Frankish, A. et al. Comparison of GENCODE and RefSeq gene annotation and the impact of reference geneset on variant effect prediction. BMC Genomics 16, S2 (2015).

3. Wu, P.-Y., Phan, J. H. & Wang, M. D. Assessing the impact of human genome annotation choice on RNA-seq expression estimates. BMC Bioinformatics 14, S8 (2013).

4. Chisanga, D., Liao, Y. & Shi, W. Impact of gene annotation choice on the quantification of RNA-seq data. BMC Bioinformatics 23, 107 (2022).

5. Zhao, S. & Zhang, B. A comprehensive evaluation of ensembl, RefSeq, and UCSC annotations in the context of RNA-seq read mapping and gene quantification. BMC Genomics 16, 97 (2015).

6. Wu, P.-Y., Phan, J. H. & Wang, M. D. The effect of human genome annotation complexity on RNA-Seq gene expression quantification. in 2012 IEEE International Conference on Bioinformatics and Biomedicine Workshops 712–717 (IEEE, 2012). doi:10.1109/BIBMW.2012.6470224.

7. Hamaguchi, Y., Zeng, C. & Hamada, M. Impact of human gene annotations on RNA-seq differential expression analysis. BMC Genomics 22, 730 (2021).

8. Chen, G. et al. Incorporating the human gene annotations in different databases significantly improved transcriptomic and genetic analyses. RNA 19, 479–489 (2013).

9. Church, D. M. et al. Modernizing Reference Genome Assemblies. PLoS Biol. 9, e1001091 (2011).

10. Guo, Y. et al. Improvements and impacts of GRCh38 human reference on high throughput sequencing data analysis. Genomics 109, 83–90 (2017).

11. Lansdon, L. A. et al. Factors Affecting Migration to GRCh38 in Laboratories Performing Clinical Next-Generation Sequencing. J. Mol. Diagn. 23, 651–657 (2021).

12. Maddirevula, S. et al. Analysis of transcript-deleterious variants in Mendelian disorders: implications for RNA-based diagnostics. Genome Biol. 21, 145 (2020).

13. Oquendo, C. J. et al. RNA sequencing uplifts diagnostic rate in undiagnosed rare disease patients. 2023.07.05.23292254 Preprint at 10.1101/2023.07.05.23292254 (2023).

14. Kremer, L. S., Wortmann, S. B. & Prokisch, H. “Transcriptomics”: molecular diagnosis of inborn errors of metabolism via RNA-sequencing. J. Inherit. Metab. Dis. 41, 525–532 (2018).

15. Frésard, L. et al. Identification of rare-disease genes using blood transcriptome sequencing and large control cohorts. Nat. Med. 25, 911–919 (2019).

16. Kremer, L. S. et al. Genetic diagnosis of Mendelian disorders via RNA sequencing. Nat. Commun. 8, 15824 (2017).

17. Mertes, C. et al. Detection of aberrant splicing events in RNA-seq data using FRASER. Nat. Commun. 12, 529 (2021).

18. Murdock, D. R. et al. Transcriptome-directed analysis for Mendelian disease diagnosis overcomes limitations of conventional genomic testing. J. Clin. Invest. 131, e141500 (2021).

19. Yépez, V. A. et al. Detection of aberrant gene expression events in RNA sequencing data. Nat. Protoc. 16, 1276–1296 (2021).

20. Yépez, V. A. et al. Clinical implementation of RNA sequencing for Mendelian disease diagnostics. Genome Med. 14, 38 (2022).

21. Lee, H. et al. Diagnostic utility of transcriptome sequencing for rare Mendelian diseases. Genet. Med. 22, 490–499 (2020).

22. Cummings, B. B. et al. Improving genetic diagnosis in Mendelian disease with transcriptome sequencing. Sci. Transl. Med. 9, eaal5209 (2017).

23. Youssefian, L. et al. Whole-Transcriptome Analysis by RNA Sequencing for Genetic Diagnosis of Mendelian Skin Disorders in the Context of Consanguinity. Clin. Chem. 67, 876–888 (2021).

24. Rentas, S. et al. Diagnosing Cornelia de Lange syndrome and related neurodevelopmental disorders using RNA sequencing. Genet. Med. 22, 927–936 (2020).

25. Gonorazky, H. D. et al. Expanding the Boundaries of RNA Sequencing as a Diagnostic Tool for Rare Mendelian Disease. Am. J. Hum. Genet. 104, 466–483 (2019).

26. Bournazos, A. M. et al. Standardized practices for RNA diagnostics using clinically accessible specimens reclassifies 75% of putative splicing variants. Genet. Med. 24, 130–145 (2022).

27. Dekker, J. et al. Web-accessible application for identifying pathogenic transcripts with RNA-seq: Increased sensitivity in diagnosis of neurodevelopmental disorders. Am. J. Hum. Genet. 110, 251–272 (2023).

28. Nurk, S. et al. The complete sequence of a human genome. Science 376, 44–53 (2022).

29. Ormond, C., Ryan, N. M., Corvin, A. & Heron, E. A. Converting single nucleotide variants between genome builds: from cautionary tale to solution. Brief. Bioinform. 22, bbab069 (2021).

30. Li, H. et al. Exome variant discrepancies due to reference-genome differences. Am. J. Hum. Genet. 108, 1239–1250 (2021).

31. Pan, B. et al. Similarities and differences between variants called with human reference genome HG19 or HG38. BMC Bioinformatics 20, 101 (2019).

32. Gao, G. F. et al. Before and After: Comparison of Legacy and Harmonized TCGA Genomic Data Commons’ Data. Cell Syst. 9, 24–34.e10 (2019).

33. Dobin, A. et al. STAR: ultrafast universal RNA-seq aligner. Bioinformatics 29, 15–21 (2013).

34. Burset, M. Analysis of canonical and non-canonical splice sites in mammalian genomes. Nucleic Acids Res. 28, 4364–4375 (2000).

35. Winkler, W. E. String Comparator Metrics and Enhanced Decision Rules in the Fellegi-Sunter Model of Record Linkage. https://eric.ed.gov/?id=ED325505 (1990).

36. Park, J., Yhim, H.-Y., Kang, K. P., Bae, T. W. & Cho, Y. G. Copy number variation analysis using next-generation sequencing identifies the CFHR3/CFHR1 deletion in atypical hemolytic uremic syndrome: a case report. Hematology 27, 603–608 (2022).

37. Zipfel, P. F. et al. Deletion of Complement Factor H–Related Genes CFHR1 and CFHR3 Is Associated with Atypical Hemolytic Uremic Syndrome. PLOS Genet. 3, e41 (2007).

38. Hamza, A. et al. The absence of CFHR3 and CFHR1 genes from the T2T-CHM13 assembly can limit the molecular diagnosis of complement-related diseases. Eur. J. Hum. Genet. 1–3 (2023) doi:10.1038/s41431-023-01350-8.

39. Hansen, J. et al. De Novo Mutations in SIK1 Cause a Spectrum of Developmental Epilepsies. Am. J. Hum. Genet. 96, 682–690 (2015).

40. Hartono, A. B. et al. Salt-Inducible Kinase 1 is a potential therapeutic target in Desmoplastic Small Round Cell Tumor. Oncogenesis 11, 1–10 (2022).

41. Ritchie, M. E. et al. limma powers differential expression analyses for RNA-sequencing and microarray studies. Nucleic Acids Res. 43, e47–e47 (2015).

42. Hoffman, G. E. & Roussos, P. Dream: powerful differential expression analysis for repeated measures designs. Bioinformatics 37, 192–201 (2020).

43. Schneider, V. A. et al. Evaluation of GRCh38 and de novo haploid genome assemblies demonstrates the enduring quality of the reference assembly. Genome Res. 27, 849–864 (2017).

44. Sondka, Z. et al. COSMIC: a curated database of somatic variants and clinical data for cancer. Nucleic Acids Res. gkad986 (2023) doi:10.1093/nar/gkad986.

45. Wadugu, B. A., et al. *U2af1* is a haplo-essential gene required for hematopoietic cancer cell survival in mice. J. Clin. Invest. 131, (2021).

46. Shirai, C. L. et al. Mutant U2AF1-expressing cells are sensitive to pharmacological modulation of the spliceosome. Nat. Commun. 8, 14060 (2017).

47. Tang, D., Zhao, X., Zhang, L. & Wang, C. Comprehensive analysis of pseudogene HSPB1P1 and its potential roles in hepatocellular carcinoma. J. Cell. Physiol. 235, 6515–6527 (2020).

48. Li, Y. I. et al. Annotation-free quantification of RNA splicing using LeafCutter. Nat. Genet. 50, 151–158 (2018).

49. Zhao, M., et al. Phen2Gene: rapid phenotype-driven gene prioritization for rare diseases. NAR Genomics Bioinforma. 2, lqaa032 (2020).

50. Postel, M. D., Culver, J. O., Ricker, C. & Craig, D. W. Transcriptome analysis provides critical answers to the “variants of uncertain significance” conundrum. Hum. Mutat. 43, 1590–1608 (2022).

51. Truty, R. et al. Spectrum of splicing variants in disease genes and the ability of RNA analysis to reduce uncertainty in clinical interpretation. Am. J. Hum. Genet. 108, 696–708 (2021).

52. Byron, S. A., Van Keuren-Jensen, K. R., Engelthaler, D. M., Carpten, J. D. & Craig, D. W. Translating RNA sequencing into clinical diagnostics: opportunities and challenges. Nat. Rev. Genet. 17, 257–271 (2016).

53. Karam, R. et al. Assessment of Diagnostic Outcomes of RNA Genetic Testing for Hereditary Cancer. *JAMA Netw*. Open 2, e1913900 (2019).

54. Aganezov, S. et al. A complete reference genome improves analysis of human genetic variation. Science 376, eabl3533 (2022).

55. Vollger, M. R. et al. Segmental duplications and their variation in a complete human genome. Science 376, eabj6965 (2022).

56. Liao, W.-W. et al. A draft human pangenome reference. Nature 617, 312–324 (2023).

57. Wang, T. et al. The Human Pangenome Project: a global resource to map genomic diversity. Nature 604, 437–446 (2022).

58. Behera, S. et al. FixItFelix: improving genomic analysis by fixing reference errors. Genome Biol. 24, 31 (2023).

59. Amar, David et al. Temporal dynamics of the multi-omic response to endurance exercise training across tissues. 2022.09.21.508770 Preprint at 10.1101/2022.09.21.508770 (2023).

60. Frankish, A. et al. GENCODE 2021. Nucleic Acids Res. 49, D916–D923 (2021).

61. Pertea, G. & Pertea, M. GFF Utilities: GffRead and GffCompare. F1000Research 9, ISCB Comm J-304 (2020).

62. Li, B. & Dewey, C. N. RSEM: accurate transcript quantification from RNA-Seq data with or without a reference genome. 16 (2011).

63. Illumina. bcl2fastq.

64. Martin, M. Cutadapt removes adapter sequences from high-throughput sequencing reads. EMBnet.journal 17, 10–12 (2011).

65. Broad Institute. Picard Toolkit. (2019).

66. Danecek, P. et al. Twelve years of SAMtools and BCFtools. GigaScience 10, giab008 (2021).

67. Cotto, K. C., et al. RegTools: Integrated analysis of genomic and transcriptomic data for the discovery of splicing variants in cancer. http://biorxiv.org/lookup/doi/10.1101/436634 (2018) doi:10.1101/436634.

68. Jenkinson, G. et al. LeafCutterMD: an algorithm for outlier splicing detection in rare diseases. Bioinformatics (2020) doi:10.1093/bioinformatics/btaa259.

69. Quinlan, A. R. & Hall, I. M. BEDTools: a flexible suite of utilities for comparing genomic features. Bioinformatics 26, 841–842 (2010).

70. Diekhans, M. gencode-backmap. (2023).

71. Hoyt, S. J. et al. From telomere to telomere: The transcriptional and epigenetic state of human repeat elements. Science 376, eabk3112 (2022).

72. Sariyar, M. & Borg, A. The RecordLinkage Package: Detecting Errors in Data. R J. 2, 61 (2010).

73. McKusick, V. A. Mendelian Inheritance in Man and Its Online Version, OMIM. Am. J. Hum. Genet. 80, 588–604 (2007).

74. Tate, J. G. et al. COSMIC: the Catalogue Of Somatic Mutations In Cancer. Nucleic Acids Res. 47, D941–D947 (2019).

75. Ghoussaini, M. et al. Open Targets Genetics: systematic identification of trait-associated genes using large-scale genetics and functional genomics. Nucleic Acids Res. 49, D1311–D1320 (2021).

76. Landrum, M. J. et al. ClinVar: improving access to variant interpretations and supporting evidence. Nucleic Acids Res. 46, D1062–D1067 (2018).

77. Amemiya, H. M., Kundaje, A. & Boyle, A. P. The ENCODE Blacklist: Identification of Problematic Regions of the Genome. Sci. Rep. 9, 9354 (2019).

78. Ogata, J. D. et al. excluderanges: exclusion sets for T2T-CHM13, GRCm39, and other genome assemblies. Bioinformatics 39, btad198 (2023).

79. Dozmorov, Mikhail et al. excluderanges.

80. Human genome issues - Genome Reference Consortium. https://www.ncbi.nlm.nih.gov/grc/human/issues.

81. Kent, W. J., Zweig, A. S., Barber, G., Hinrichs, A. S. & Karolchik, D. BigWig and BigBed: enabling browsing of large distributed datasets. Bioinformatics 26, 2204–2207 (2010).

82. Mc Cartney, A. M., et al. Chasing perfection: validation and polishing strategies for telomere-to-telomere genome assemblies. Nat. Methods 19, 687–695 (2022).

83. Kent, W. J. et al. The Human Genome Browser at UCSC. Genome Res. 12, 996–1006 (2002).

84. Karolchik, D. et al. The UCSC Table Browser data retrieval tool. Nucleic Acids Res. 32, D493–D496 (2004).

85. Nassar, L. R. et al. The UCSC Genome Browser database: 2023 update. Nucleic Acids Res. 51, D1188–D1195 (2023).

