## Supplemental Figures for "Impact of genome build on RNA-seq interpretation and diagnostics"

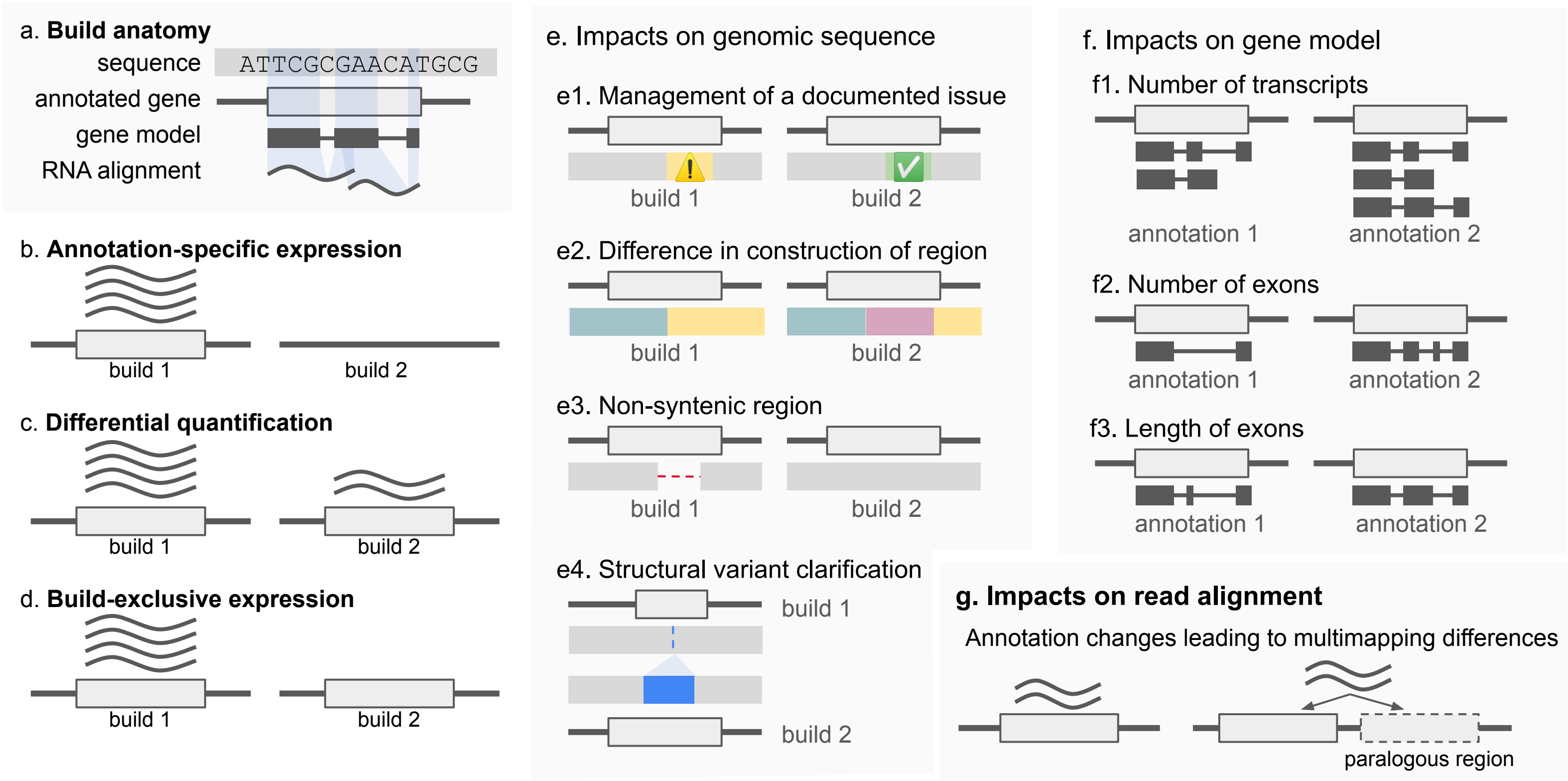


**Supplemental Figure 1: Potential differences between builds**
Examples of how different builds can cause transcriptomic differences. **a,** Builds can have a difference due to underlying sequence, the gene annotation, the gene model, and how the RNA aligns. There are three ways in which expression can differ: **b**, annotation-specific expression in which a gene is only annotated in one build, **c**, build-dependent expression in which a gene is annotated and in both builds but expressed at different levels, and **d,** build-exclusive expression in which genes are annotated in both build but only expressed in one of them. **e,** A genome sequence can change between builds due to **e1**, an issue being resolved, **e2**, the region being constructed differently, **e3**, presence of a novel region, or **e4**, a structural variant being clarified. **f**, A gene model can change between builds due to **f1**, the number of transcripts, **f2**, the number of exons, **f3**, or the length of exons varying. **g**, One way in which read alignment can vary is due to multimapping differences.

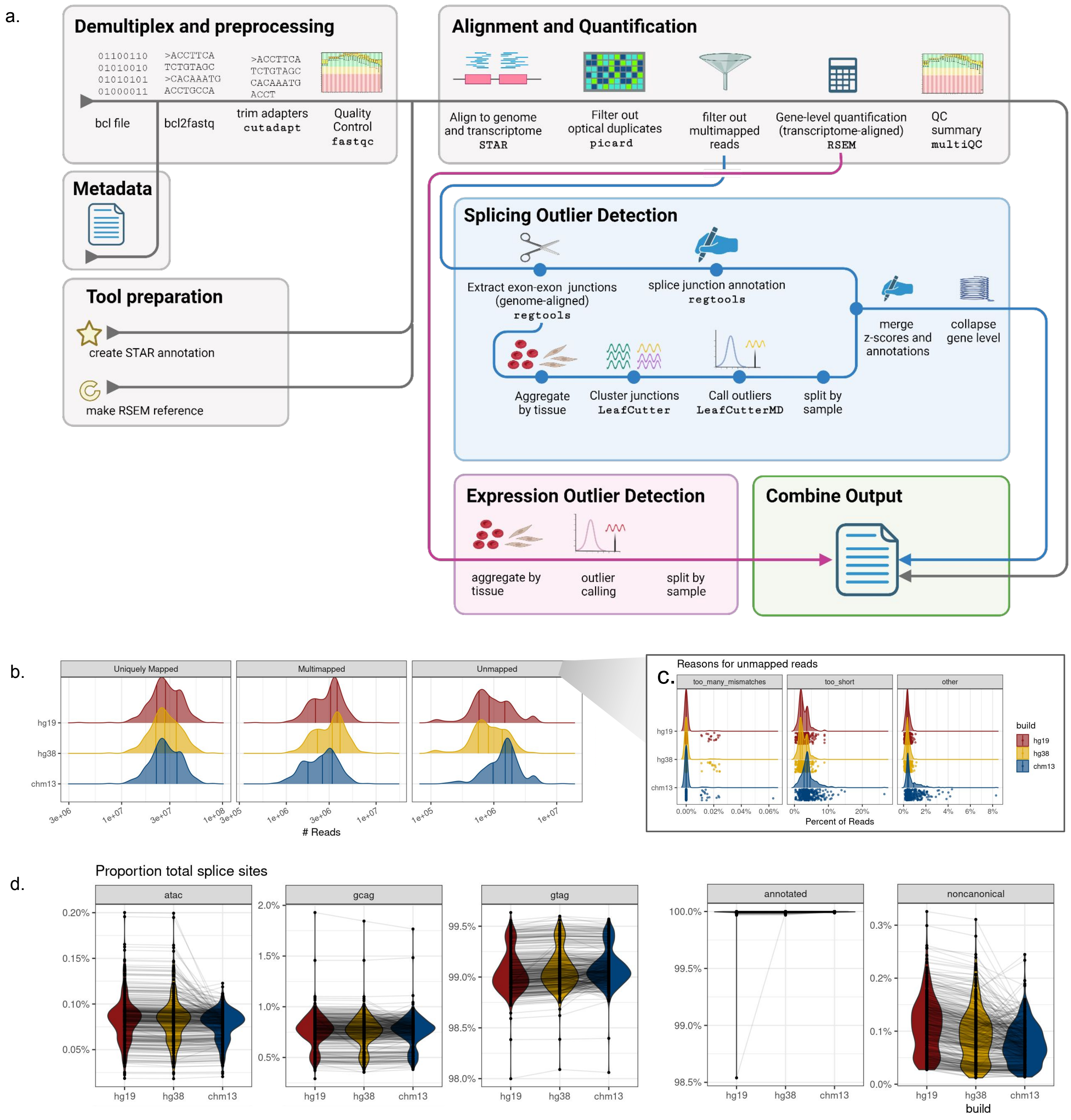


**Supplementary Figure 2: Pipeline and QC**

**a**, Schematic of the pipeline used to generate data. See https://github.com/raungar/build_rnaseq_paper_public for more details. **b**, Distribution of reads that were uniquely mapped, multi-mapped, and unmapped across builds. **c**, Explanations for unmapped reads. **d**, Proportion of splice sites were canonical, annotated, and noncanonical.

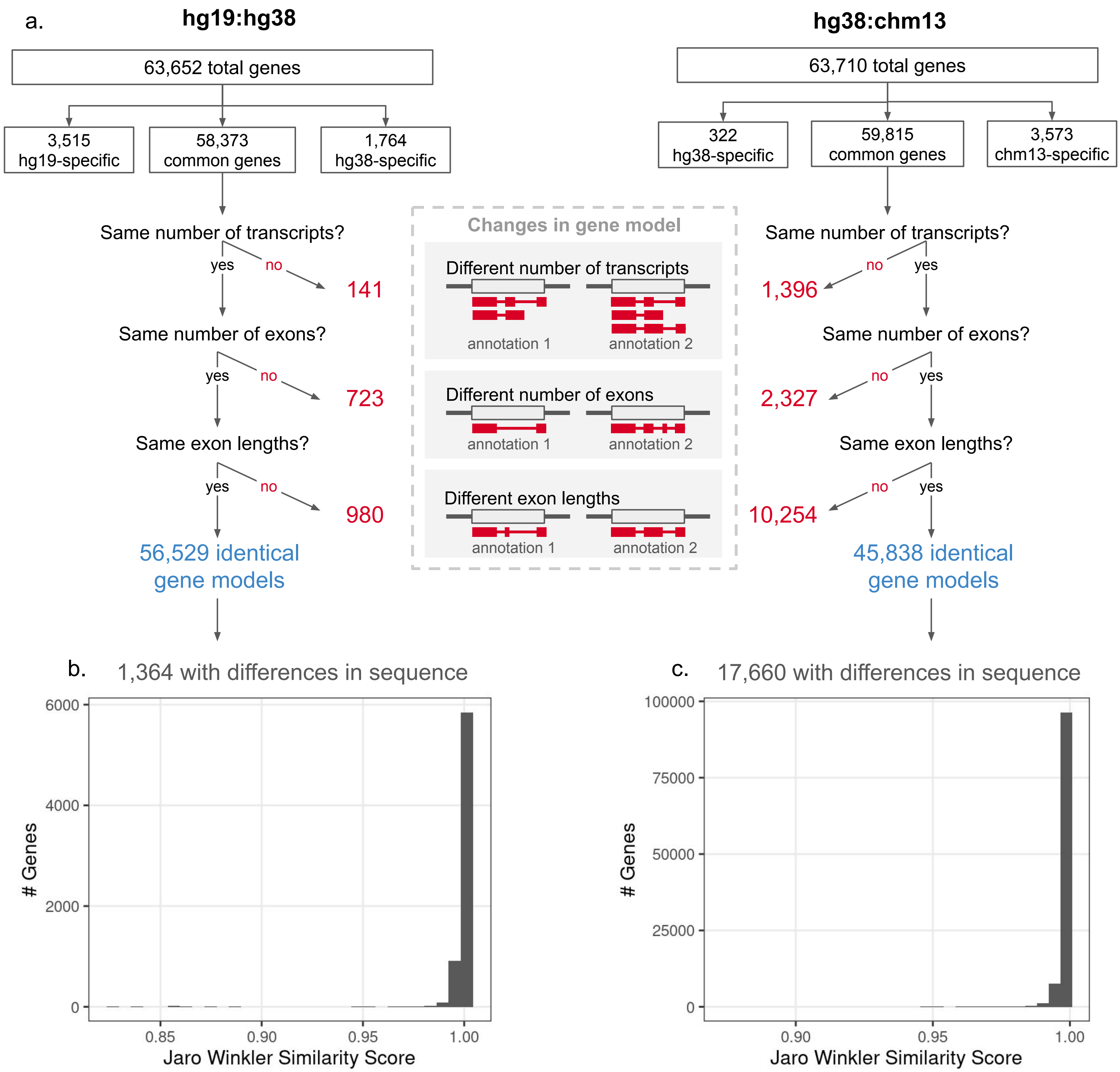


**Supplementary Figure 3: Gene model comparison flowchart**

**a**, Flowchart logic map for gene annotation similarity characterization. For genes with differences in sequence, the distribution of Jaro-Winkler similarity score is plotted for (**b**) hg19:hg38 and (**c**) hg38:chm13.

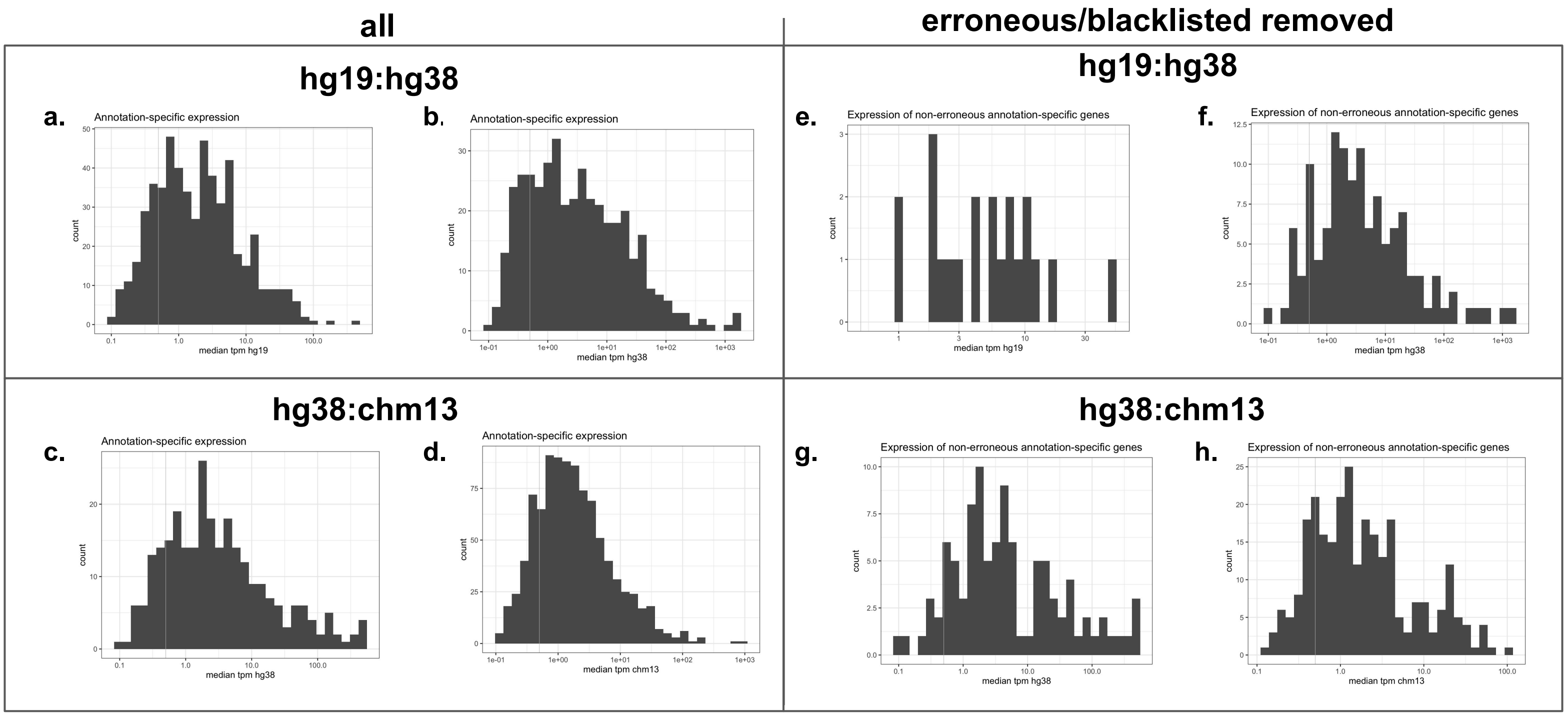


**Supplementary Figure 4: Distribution of annotation-specific expression**
For the hg19:hg38 comparison, the distribution of median TPM for **a,** hg19 annotation-specific genes and **b,** hg38 annotation-specific genes; for the hg38:chm13 comparison the **c**, hg38 annotation-specific genes and **d**, chm13 annotation-specific genes. The right side represents the distribution of the median TPM when genes in erroneous or blacklisted regions were removed for the hg19:hg38 comparison **e,** hg19 annotation-specific genes, **f,** hg38 annotation-specific genes; for the hg38:chm13 comparison the **g,** hg38-specific genes and **h,** chm13-specific genes

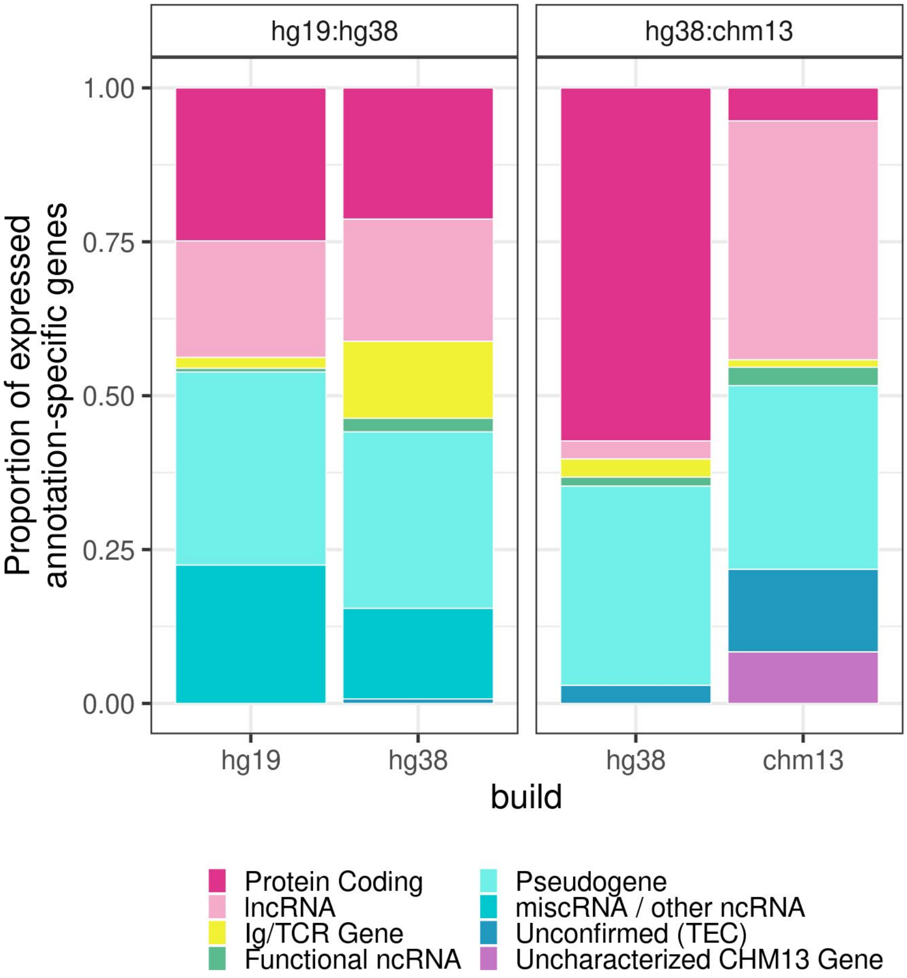


**Supplementary Figure 5: Biotypes of annotation-specific genes**
Distribution of biotypes (colored) of annotation-specific genes for the hg19:hg38 and hg38:chm13 comparison.

a.

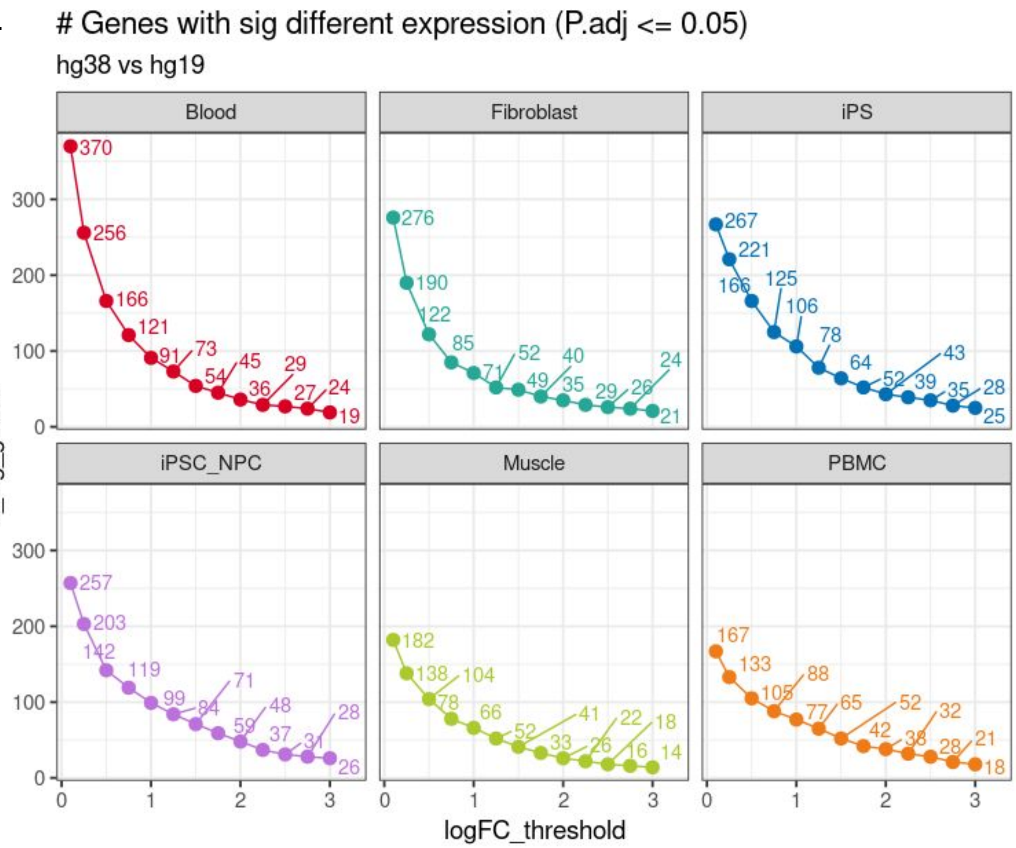


b.

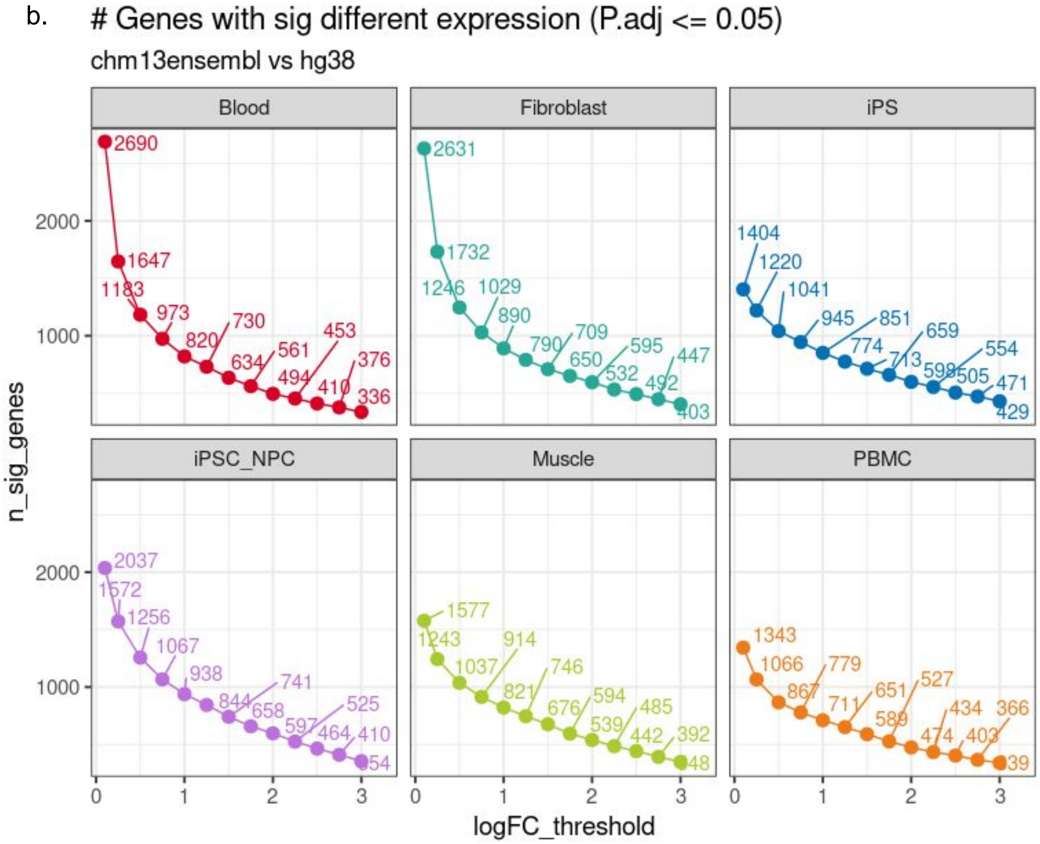


**Supplementary Figure 6: Differential quantification across logFC thresholds**
The number of genes detected at different logFC cutoffs across tissue types for the **a,** hg19:hg38 and **b,** hg38:chm13 comparisons.

a.

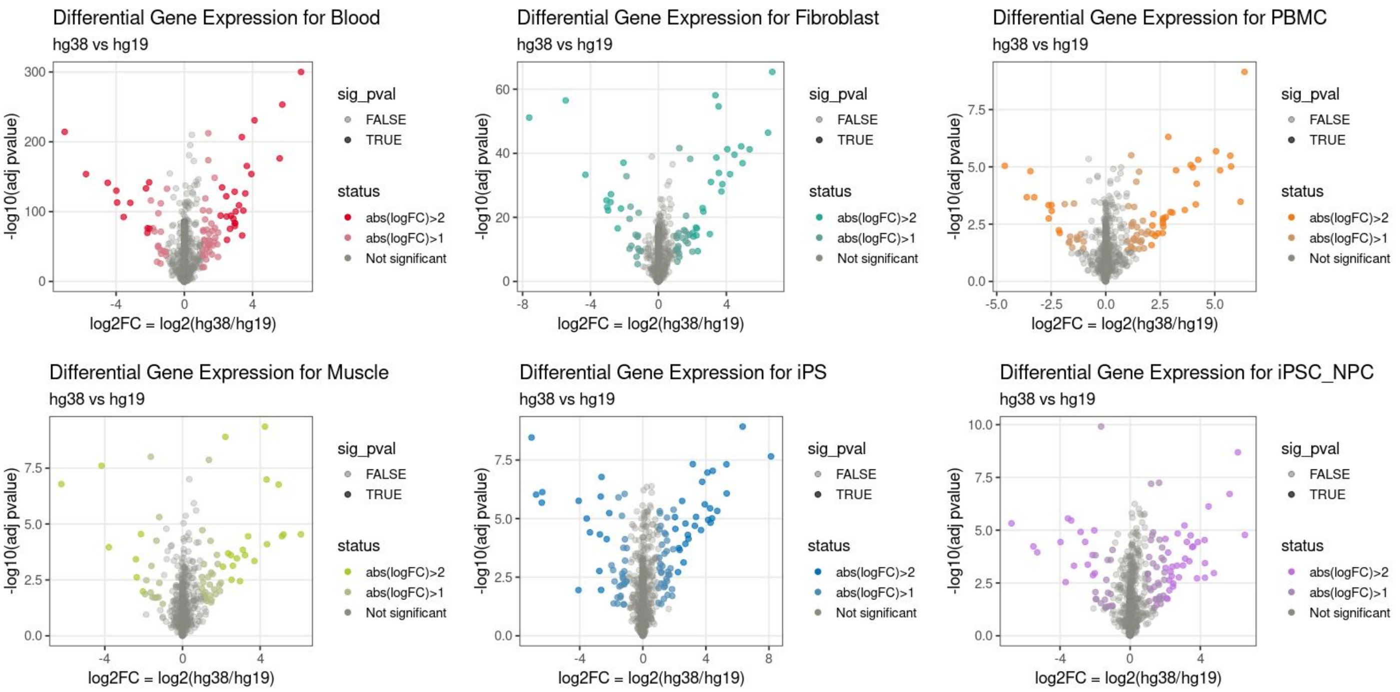


b.

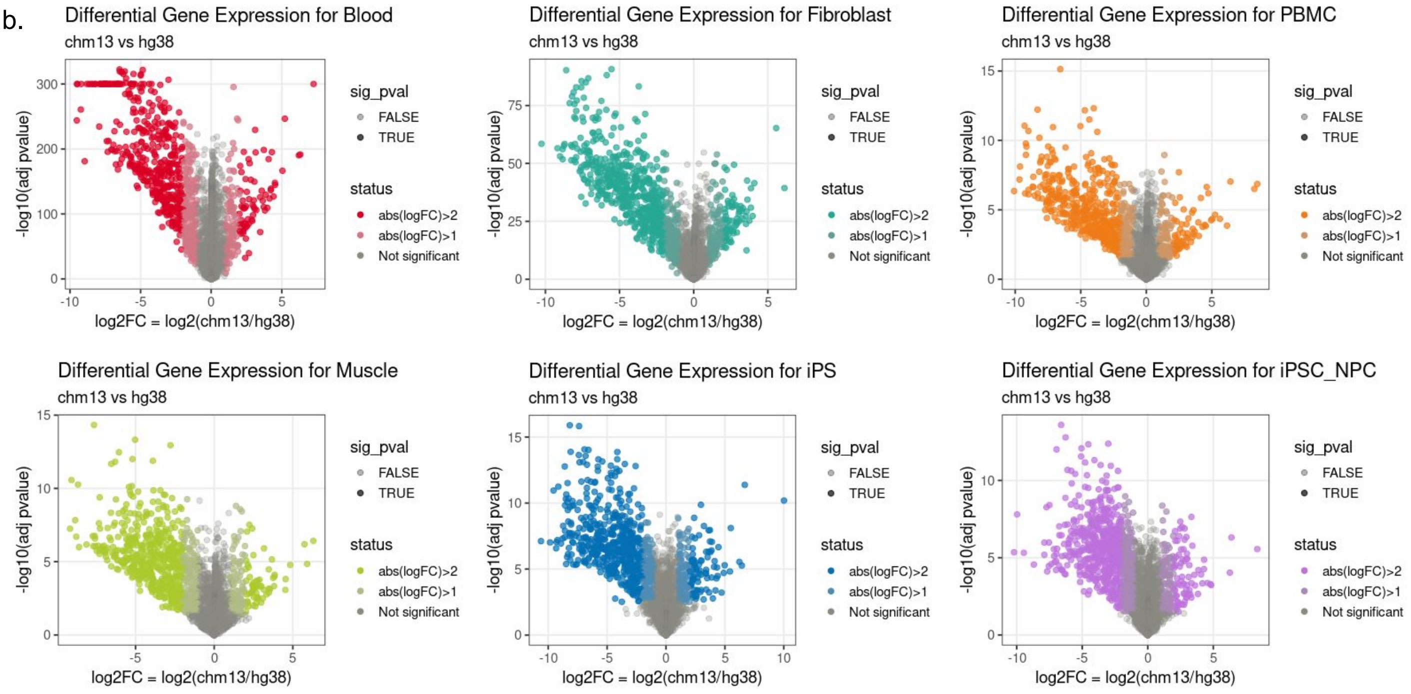


**Supplementary Figure 7: Differential quantification across tissues**
Volcano plots displaying the significance (-log10(p-value), y-axis) and magnitude (logFC, x-axis) of genes tested in each tissue for **a,** the hg19 vs hg38 comparison and **b,** the hg38 vs chm13 comparison.

**a.**

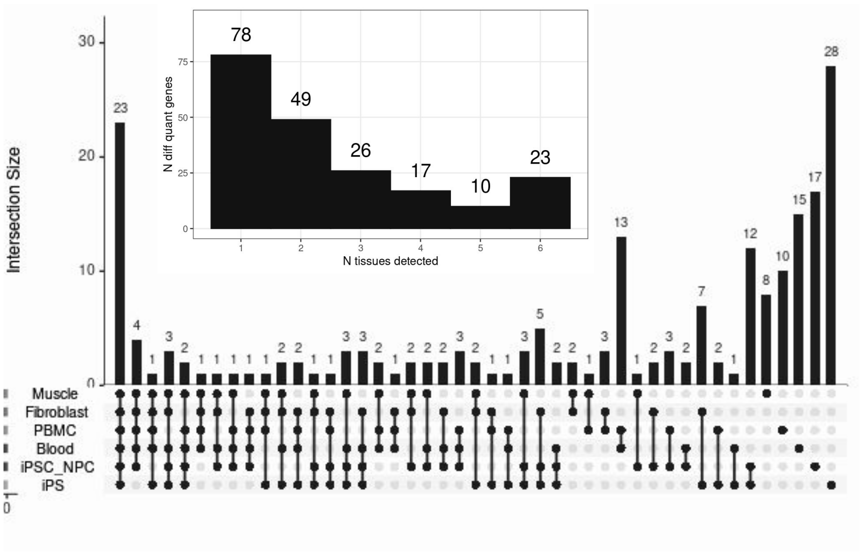


**b**

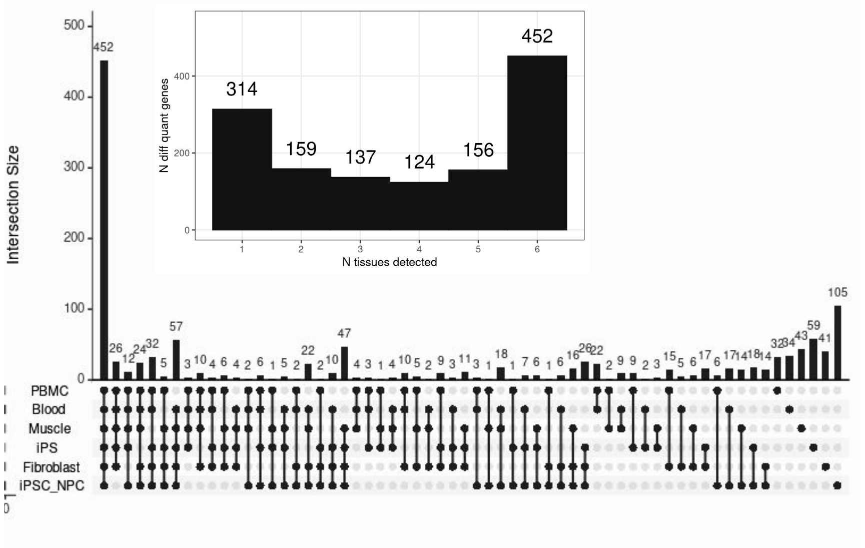


**Supplementary Figure 8: Differential quantification sharing across tissues**
Sharing of differentially quantified genes across tissues for the **a**, hg19:hg38 comparison and **b**, hg38:chm13 comparison. Insets show the number of differentially quantified genes detected in 1 to 6 biospecimen types.

a.

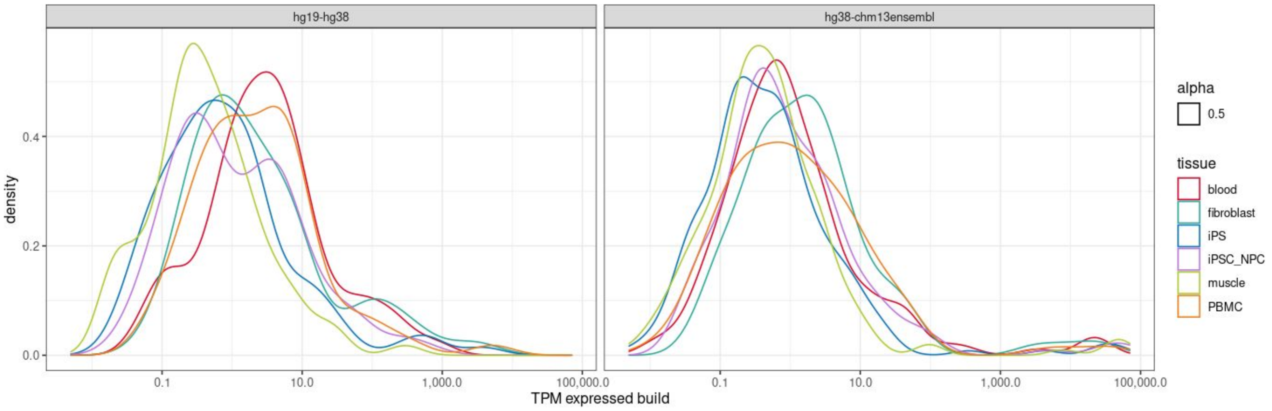


b

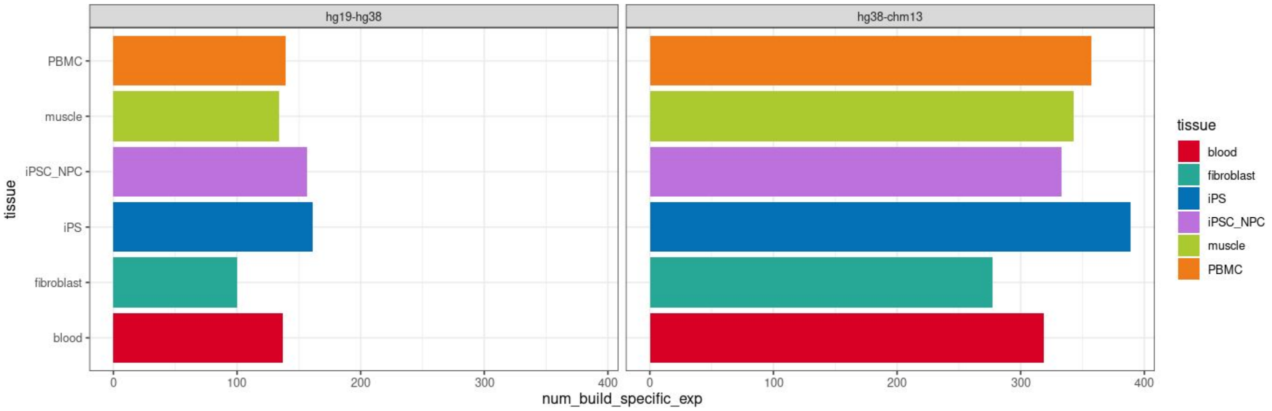


**Supplementary Figure 9: Build-exclusive gene expression**

**a.** The distribution of the median TPM of genes that were expressed in one build exclusively and **b**, Number of genes considered to exhibit build-exclusive expression across tissues.

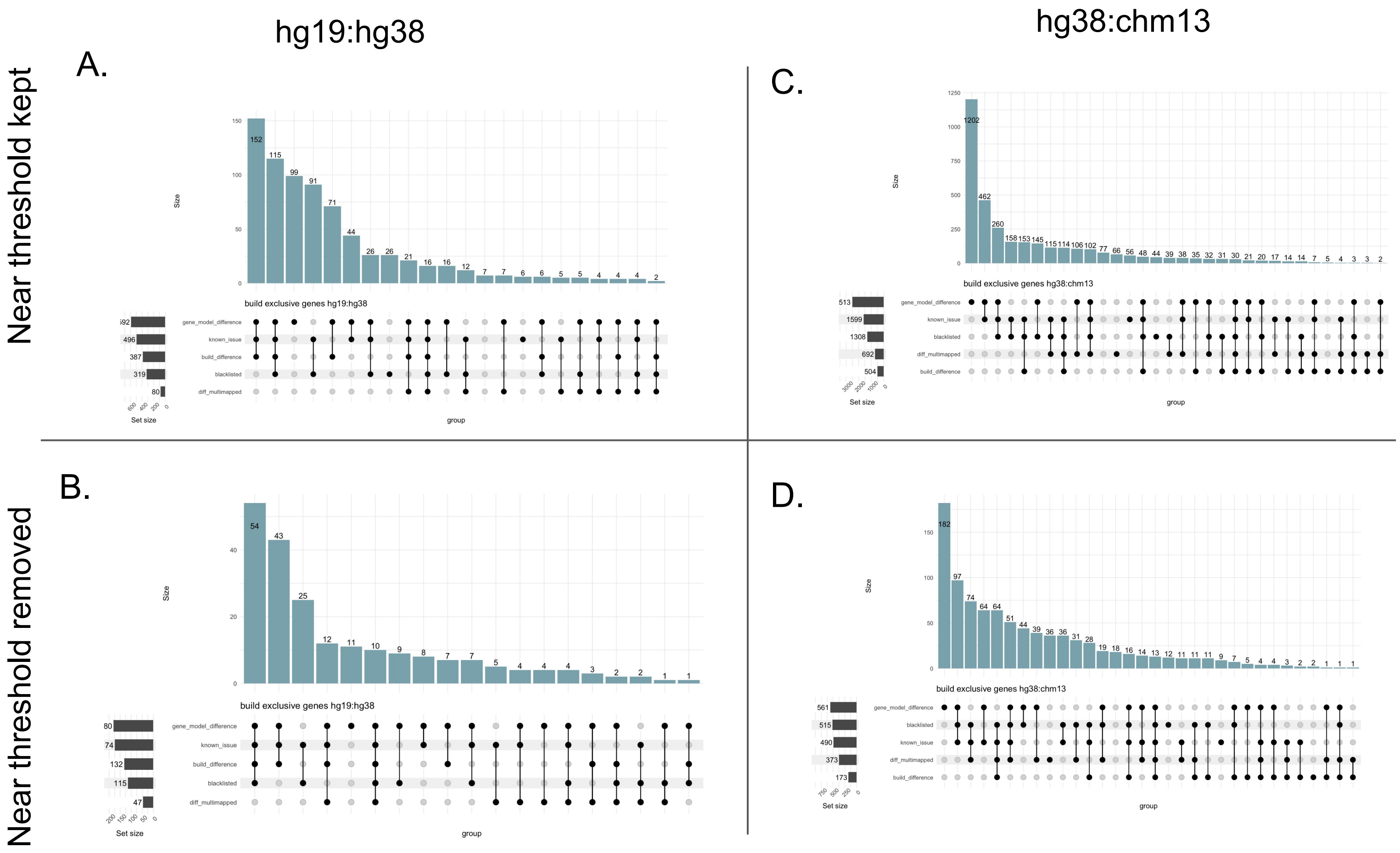


**Supplementary Figure 10. Build-exclusive expression reasons**
The reasons as to why a gene likely had expression exclusive to a given build for **a,** hg19:hg38 and **b,** with genes near the threshold of being defined as build-exclusive is removed; and for **c,** hg38:chm13 build-exclusive genes and with **d,** genes near the threshold are removed**.**

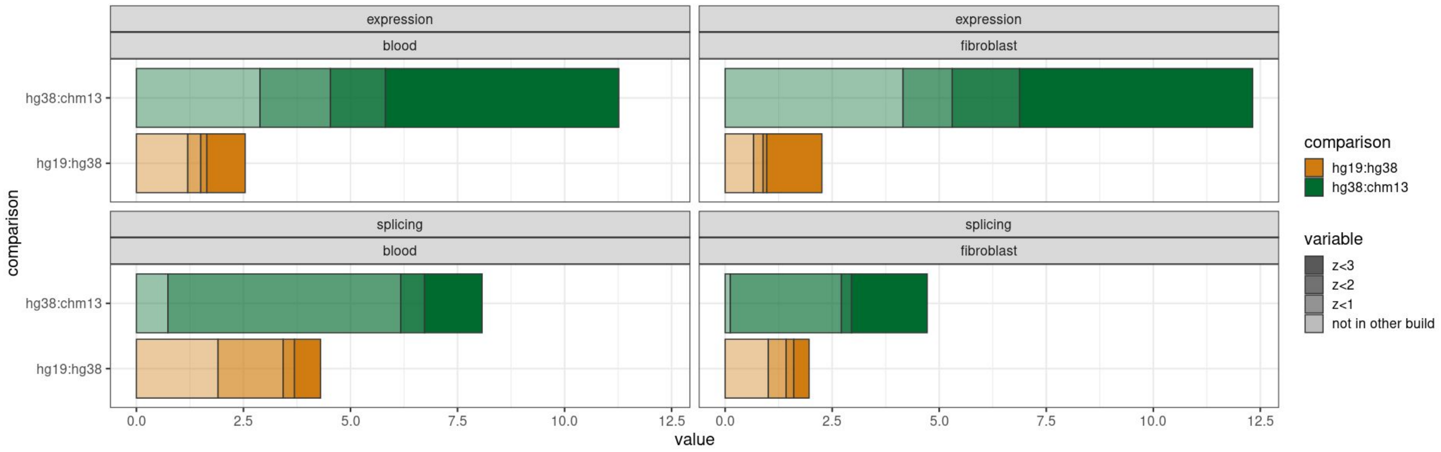


**Supplementary Figure 11: Outlier consistency comparisons (zoomed in)**
This plot shows Figure 5c zoomed in. Expression outlier consistency between hg19:hg38 (left) and hg38:chm13 (right). In orange the outliers that are consistent between hg19 and hg38, and in dark green the number of outliers consistent between hg38 and chm13. In lighter shades, the number of outliers with a z-score greater than 3 in chm13 but less than 3 in hg38 (or greater than 3 in hg38 but less than 3 in hg19) and so forth. The lightest shares are outlier in the reference build (ex chm13) but are NA in the comparison build (ex hg38) due to lack of quantification in that build.  This is faceted by tissue type, and expression vs splicing outliers.

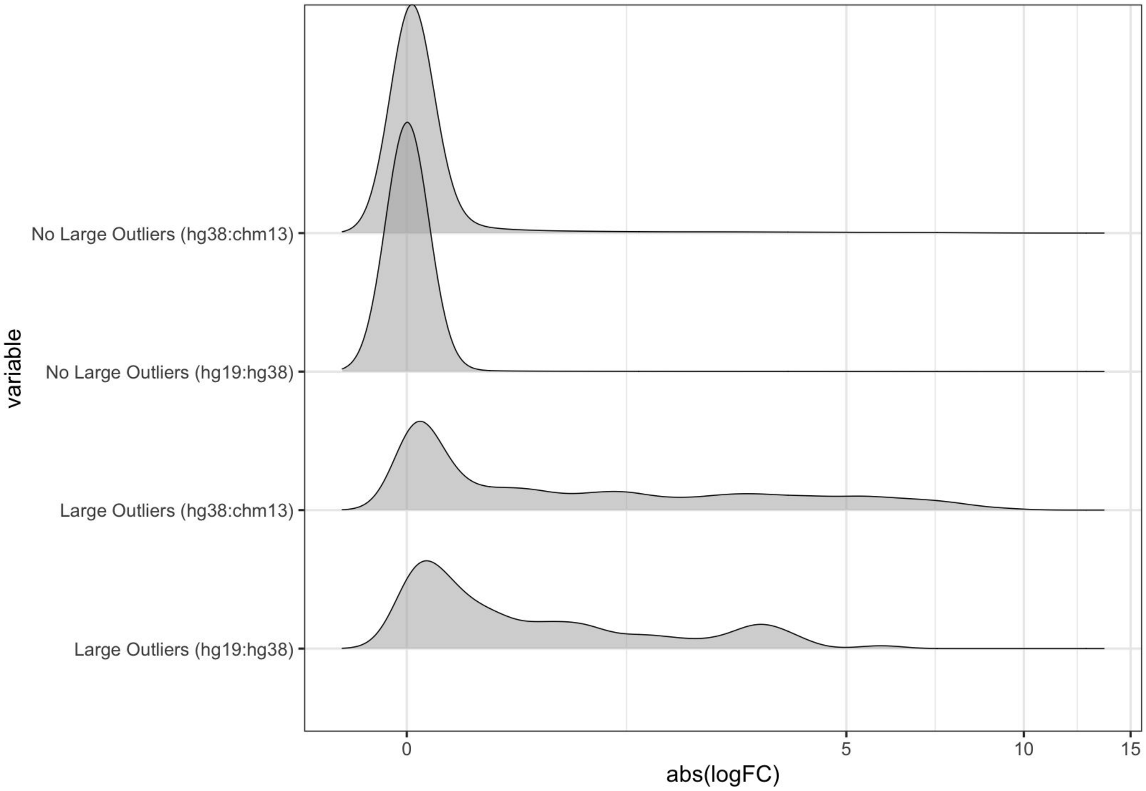


**Supplementary Figure 12: LogFC distribution stratified by large change in outlier status**
The distribution of the differential quantification logFC when stratifying by genes that had at least one large change in outlier status for a given individual.
